# Safety and immunogenicity of four sequential doses of NVX-CoV2373 in adults and adolescents: a phase 3, randomized, placebo-controlled trial (PREVENT-19)

**DOI:** 10.1101/2024.11.07.24316930

**Authors:** Germán Áñez, Alice McGarry, Wayne Woo, Karen L. Kotloff, Cynthia L. Gay, Mingzhu Zhu, Shane Cloney-Clark, Joy Nelson, Haoua Dunbar, Miranda R. Cai, Iksung Cho, Zhaohui Cai, Raj Kalkeri, Joyce S. Plested, Nita Patel, Katherine Smith, Anthony M. Marchese, Gregory M. Glenn, Raburn M. Mallory, Lisa M. Dunkle, the 2019CoVn-301 Study Investigators

**Affiliations:** Novavax, Inc., Gaithersburg, Maryland, USA; Center for Vaccine Development and Global Health, University of Maryland School of Medicine, Baltimore, Maryland, USA; Institute for Global Health and Infectious Diseases, University of North Carolina School of Medicine, Chapel Hill, North Carolina, USA

**Keywords:** COVID-19, vaccine, durability, reactogenicity, booster, adolescent

## Abstract

**Background:** NVX-CoV2373, a recombinant SARS-CoV-2 spike (rS) protein vaccine with Matrix-M™ adjuvant, has been authorized for use in adults and adolescents. PREVENT-19 (NCT04611802/2019nCoV-301), a pivotal phase 3, randomized, placebo-controlled trial demonstrated robust efficacy of a primary, 2-dose series of NVX-CoV2373 against COVID-19.

**Methods:** Protocol expansions to PREVENT-19 included enrollment of adolescents (aged 12 to <18 years) and administration of 3^rd^ and 4^th^ doses of NVX-CoV2373 to adults and adolescents. Participants randomized 2:1 received NVX-CoV2373 or placebo 21 days apart; 3^rd^ and 4^th^ doses were administered ≥6 months after the preceding dose. Secondary and additional assessments included post–3^rd^- and 4^th^-dose immune responses (neutralizing antibody [nAb], anti-rS IgG, human angiotensin-converting enzyme-2–receptor binding inhibition [hACE2-RBI]) and response durability (post–3^rd^ dose) to ancestral virus; cross-reactivity to Omicron subvariants; safety; and reactogenicity.

**Results:** Immune responses were observed against ancestral virus after two doses of NVX-CoV2373 but not after placebo. In both adults and adolescents, additional doses of NVX-CoV2373 increased nAb titers, anti-rS IgG levels, and hACE2-RBI; durable responses were recorded 8 months post 3^rd^ dose. nAb responses post 3^rd^ dose were noninferior to those post primary series. Cross-reactivity to BA.5 and BQ.1.1 variants was also observed, with anti-rS IgG levels post 3^rd^ or 4^th^ dose exceeding previously reported correlates of protection. Additional doses of NVX-CoV2373 were well tolerated, with no new safety signals.

**Conclusions:** NVX-CoV2373 elicited robust and durable humoral immune responses to ancestral SARS-CoV-2 as a 3^rd^ and 4^th^ dose after the primary series in adults and adolescents. Cross-reactivity to relevant variants provides insight into potential protection against antigenically related, but shifted, viral strains. Additional doses of NVX-CoV2373 were well tolerated with no new safety signals. These results support the utility of this vaccine platform and continued updates, based on currently circulating strains, to help effectively combat SARS-CoV-2 infection.

**Highlights:**

- NVX-CoV2373 elicits robust and durable humoral immune responses to SARS-CoV-2
- 3^rd^ and 4^th^ subsequent homologous doses are immunogenic and well tolerated
- NVX-CoV2373 was cross-reactive to variants circulating at the time of this study

## 1. Introduction

NVX-CoV2373 is a recombinant spike (rS) protein prototype vaccine to ancestral SARS-CoV-2 with a saponin-based adjuvant (Matrix-M™) that was authorized for use in adults and adolescents in the United States (US), and many other regions, for primary vaccination and additional homologous or heterologous doses.^1,2^ An updated version, NVX-CoV2601, contains XBB.1.5 rS and was authorized in 2023–2024 for use in people aged ≥12 years.^2,3^ In 2024, the World Health Organization and US Centers for Disease Control and Prevention estimates suggested that XBB.1.5-based COVID-19 vaccines were effective against symptomatic SARS-CoV-2 infection, including cross protection.^4,5^ Based on SARS-CoV-2 variant evolution, another COVID-19 vaccine update was initiated for 2024–2025, targeting the JN.1-lineage,^6^ resulting in updated authorizations/approvals,^7^ including for the Novavax COVID-19 vaccine. Despite the availability of updated formulations, immunogenicity of repeated homologous prototype vaccinations remains relevant in the context of cross-protection among variants and durability of humoral immune responses.

Prefusion Protein Subunit Vaccine Efficacy Novavax Trial–COVID-19 (PREVENT-19), was a pivotal phase 3, randomized, placebo-controlled, multicenter clinical trial of NVX-CoV2373 conducted in ∼30,000 adults in the US and Mexico (NCT04611802/2019nCoV-301). The primary objective was met with 90% vaccine efficacy against mild, moderate, or severe COVID-19.^8^ Additional analyses demonstrated 100% efficacy against moderate-to-severe disease and hospitalization ≥7 days after the 2^nd^ dose.^8,9^ No preliminary safety concerns were raised, with a median 2-month safety follow-up.

A pediatric expansion was initiated in April 2021 to enroll adolescents (aged 12 to <18 years) in the US; preliminary safety, immunogenicity, and efficacy were demonstrated in an interim report.^10^ Additionally, to align with evolving public health recommendations, protocol amendments allowed participants who completed an NVX-CoV2373 primary series to receive a 3^rd^ dose and subsequent 4^th^ dose. This report describes immunogenicity and safety results from adults and adolescents enrolled in PREVENT-19, including outcomes from participants who received additional doses of NVX-CoV2373. Immunogenicity against the ancestral strain and cross-reactivity against viral variants predominant at the time of this report are included.

## 2. Methods

### 2.1. Study design

Enrollment criteria for adult^8^ and adolescent (aged 12 to <18 years)^10^ participants were described previously. Participants were randomized 2:1 to receive two intramuscular injections of either NVX-CoV2373 or placebo 21 days apart (**Figure S1**). After accrual of safety data to support Emergency Use Authorization, participants could opt in for the blinded crossover and receive alternate treatment from their original randomization group. Adult participants in study follow-up for ≥6 months (≥5 months for adolescents) after receipt of the NVX-CoV2373 primary series were eligible for a 3^rd^ dose (open label); a 4^th^ dose was offered at select sites ≥6 months later.

### 2.2. Objectives

Results of primary efficacy, initial safety, and some secondary objectives post primary series in adults and adolescents have been reported previously.^8,10^ Final adult safety outcomes, as well as updated immunogenicity and safety analyses post–primary series dosing and outcomes and after 3^rd^ and 4^th^ doses in adults and adolescent participants are reported here. Secondary immunogenicity objectives were neutralizing antibody (nAb) titers, serum IgG levels, and human angiotensin-converting enzyme-2 (hACE2) receptor-binding inhibition (RBI) before and 14 days after (day 35) the NVX-CoV2373 primary series. The 14-day timepoint for the primary series was used in this study based on timing in pivotal COVID-19 vaccine trials in the height of the pandemic.^8,11^ Immune responses 28 days after the 3^rd^ and 4^th^ doses and comparison of these to the primary series responses were assessed.

Safety assessment included collection of solicited local and systemic reactogenicity for 7 days after each study dose and incidence and severity of unsolicited adverse events (AEs) for 28 days after each study dose. Overall safety included frequency and severity of treatment-related medically attended AEs (MAAEs), AEs of special interest (AESI), serious AEs (SAEs), and death due to any cause throughout study follow-up.

### 2.3. Procedures

Participants received NVX-CoV2373 (5 μg SARS-CoV-2 rS protein and 50 μg Matrix-M adjuvant) via a 0.5-mL intramuscular injection. Blood samples for measurement of serum antibody responses were collected on day 0 (before the 1^st^ dose), day 35 (14 days following the 2^nd^ dose), and before and 28 days after the 3^rd^ and 4^th^ doses. Blood samples and nasal swabs were obtained from participants before the primary series, 3^rd^, and 4^th^ doses to determine serologic and virologic status (SARS-CoV-2 anti-nucleoprotein [NP] serum antibodies [Elecsys^®^ Anti-SARS-CoV-2 assay; University of Washington, Seattle, WA, USA]; SARS-CoV-2 RT-PCR [Abbott Real Time Quantitative SARS-CoV-2 assay; University of Washington, Seattle, WA, USA]).

Reactogenicity (solicited AEs) data were documented by participants via an eDiary for 7 days after dosing. Unsolicited treatment-emergent AEs (TEAEs) were collected through 28 days after each study dose; treatment-related MAAEs, AESI, SAEs, and death (any cause) were documented by study investigators through end of study.

### 2.4. Assessments

Serum was analyzed with validated assays for: nAbs specific to ancestral SARS-CoV-2 (via a microneutralization inhibitory concentration of 50% [MN_50_]; 360biolabs, Melbourne, Australia),^12^ anti-rS IgG antibody levels via enzyme-linked immunosorbent assay (ELISA) as geometric mean ELISA units (GMEUs),^13^ and hACE2 RBI antibodies to rS protein^14^ (both assays; Novavax Clinical Immunology, Gaithersburg, MD, USA). Fit-for-purpose assays were used for anti-rS IgG and nAbs against variant strains (ie, BA.5, BQ.1.1).^15^

Solicited AEs were summarized by local/systemic symptom and maximum severity. Unsolicited AEs were coded using the Medical Dictionary for Regulatory Activities (MedDRA) version 25.0. AEs were categorized by severity and relationship to the study vaccine.

### 2.5. Statistical analyses

The per-protocol immunogenicity (PP-IMM) analysis sets for each dose included participants with both a baseline (pre-dose) and ≥1 post-dose sample and no exclusionary protocol violations. Participants with RT-PCR–positive nasal swabs and/or anti-NP seropositivity for SARS-CoV-2 from baseline to day 35 and/or through 28 days after the 3^rd^ or 4^th^ study dose were excluded from PP-IMM analyses. All statistical analyses were performed using SAS^®^ version 9.4 or higher (SAS Institute, Cary, NC).

Geometric means were defined as the antilog of the mean of the log-transformed values. Geometric mean fold rise (GMFR) was defined as the ratio of post-dose to pre-dose geometric mean values within a study group. Data were summarized by trial vaccine group using 95% CI, calculated based on the *t*-distribution of the log-transformed values for geometric means or GMFR, then back-transformed to the original scale for presentation. For ratio calculations, concentration values below the lower limit of quantification (LLOQ) were replaced by 0.5 × LLOQ. Seroconversion rate (SCR) was defined as a percentage of participants with a ≥4-fold higher antibody level post versus pre-dose. The SCR 95% CIs were calculated using the exact Clopper–Pearson method.

Noninferiority of the 3^rd^-dose response to that following the primary series was assessed by comparing the nAb titers 28 days after the 3^rd^ dose with the response obtained 14 days after the primary series. To establish noninferiority, the following criteria were to be met: lower bound of two-sided 95% CI for the ratio of GMTs (GMT_third_/GMT_primary_) >1.0 and lower bound of the two-sided 95% CI for the difference of SCR (SCR_third_−SCR_primary_) >−10%. The 95% CI for SCR difference was calculated using the Tango method. Serum samples from randomly selected adult and adolescent PP-IMM subsets, who received all four doses, were used to assess immune responses against ancestral SARS-CoV-2 and variants at the specified time points.

The safety analysis sets included all participants who received ≥1 primary series study dose and were analyzed according to study material received; participants in the 3^rd^- and 4^th^-dose safety sets must have received these respective additional vaccinations. Reactogenicity data are reported for participants in the safety analysis set with ≥1 eDiary entry for the associated dose. Safety data were summarized descriptively, and no statistical tests were performed. Unsolicited AEs were reported by number and percentage, with corresponding exact 95% CI using the Clopper–Pearson method.

### 2.6. Ethical oversight

This study was performed in accordance with the International Conference on Harmonization Good Clinical Practice guidelines and overseen by the NIAID DSMB. The protocol, amendments, and overall oversight were provided by institutional review boards, research ethics committees, and the National Institute of Allergy and Infectious Disease (NIAID) Data Safety and Monitoring Board (DSMB), according to local regulations. Written informed consent was received from all participants before enrollment.

## 3. Results

### 3.1. Study populations

Adult participants (N=29,943; screened from 27 December 2020 to 18 February 2021) were randomized to receive NVX-CoV2373 (N=19,961) or placebo (N=9982); 713 and 337 participants were included in the respective PP-IMM analysis sets (**Figure 1A**). After crossover, 226 and 160 adult participants were in the 3^rd^- and 4^th^-dose sets, respectively (**Figure 1B**). Of 2247 adolescent participants, 1491 and 756, respectively, were randomized to receive NVX-CoV2373 and placebo (PP-IMM sets: 1126 and 537); there were 122 and 27 adolescent participants analyzed for immunogenicity after the 3^rd^ dose and 4^th^ dose, respectively (**Figure 1**).

**Figure 1.**
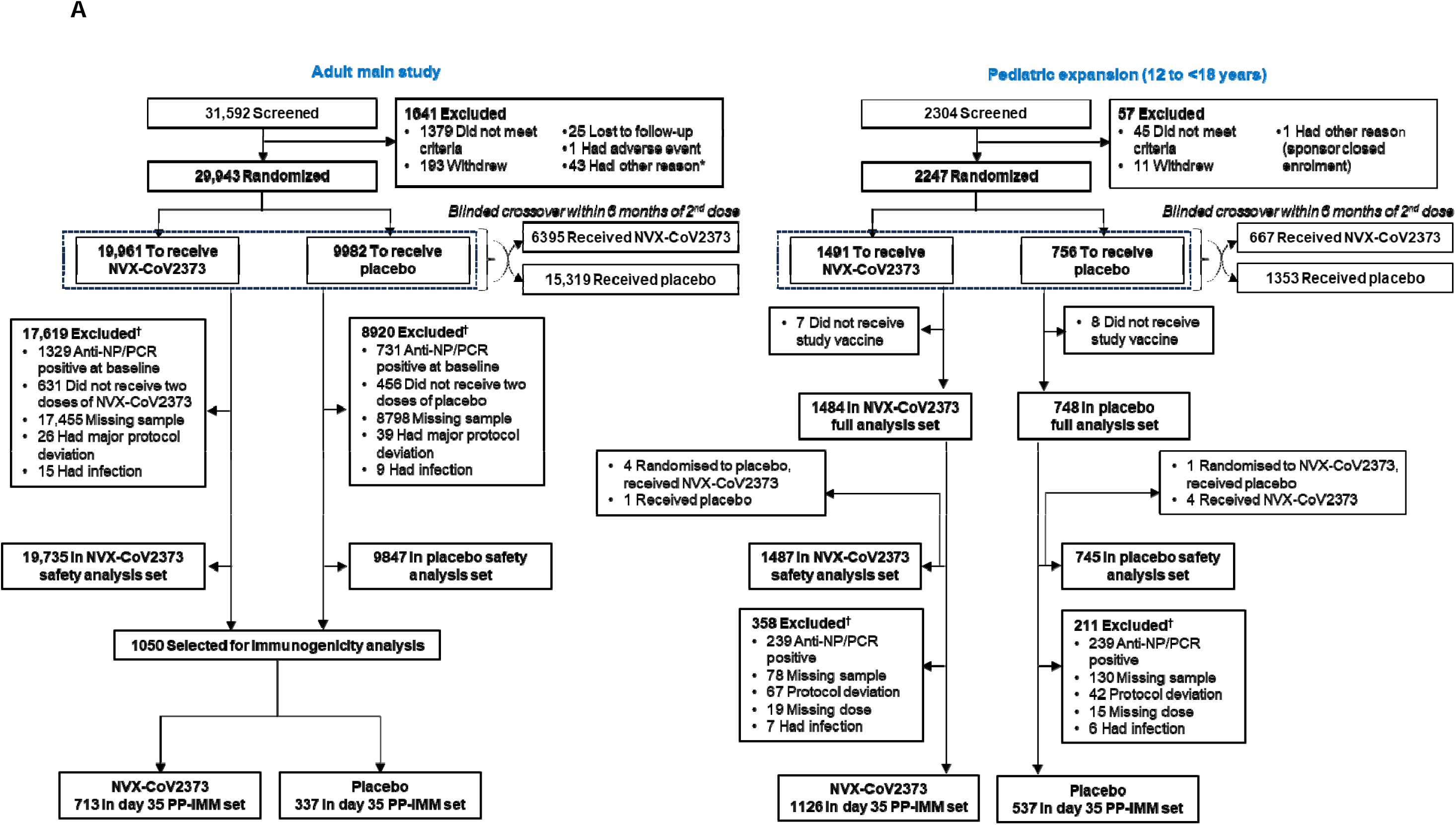
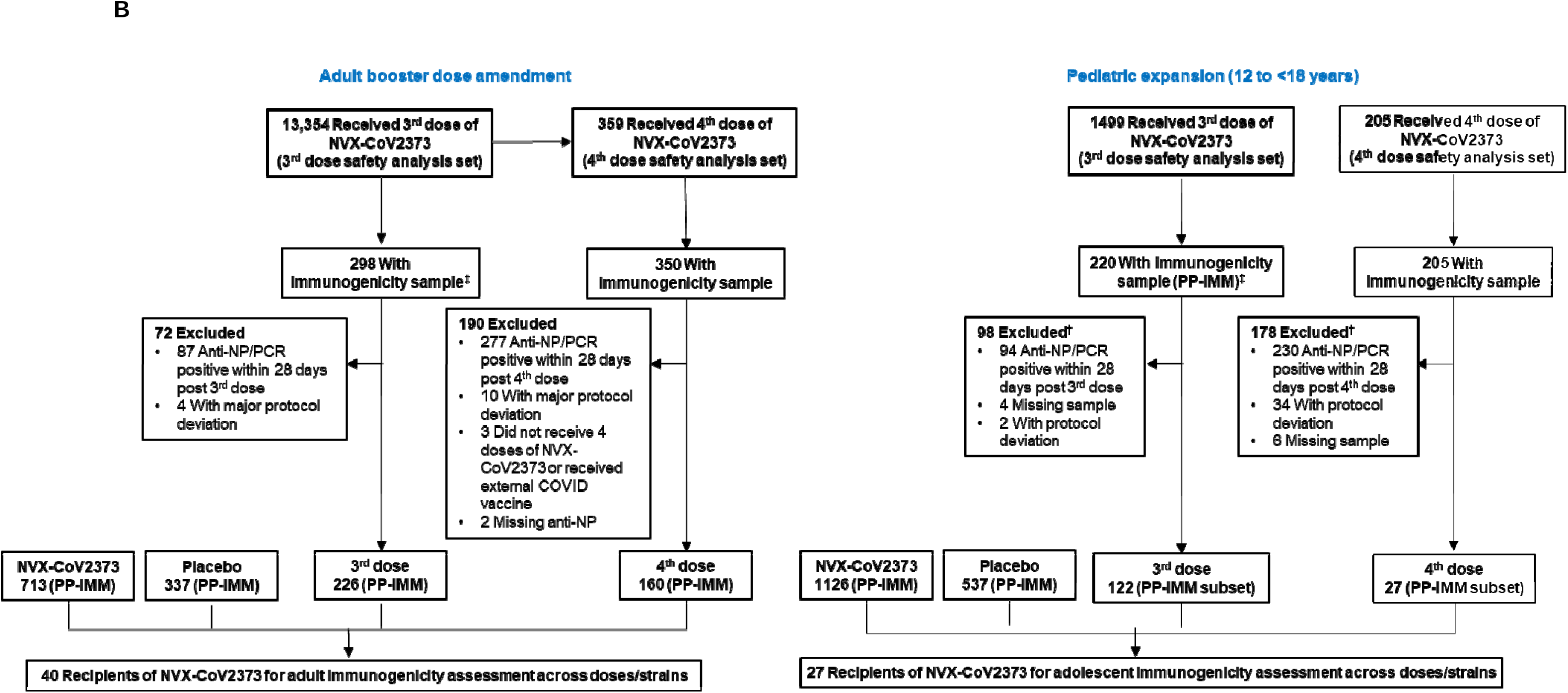
CONSORT diagram. Participant enrollment and disposition for adult and adolescent populations. (A) Primary series and crossover. (B) Dose expansion. Part of this figure was adapted from Añez G, et al. *JAMA Netw Open*. 2023;6(4):e239135. *Other reasons include receipt of another COVID-19 vaccine, noncompliance, investigator decision, incarceration, and pregnancy. ^†^Participants could have more than one reason for exclusion. ^‡^Participants with an immunogenicity sample after the 3rd dose were a combination of those who received 2 doses of NVX-CoV2373 in the initial series and those who received 2 doses during blinded crossover. ^§^Three participants were excluded from the adult 4^th^-dose safety analysis set due to receipt of an external COVID-19 vaccine or an extra dose of NVX-CoV2373; three participants excluded from the adolescent PP-IMM set who received only 3 doses of NVX-CoV2373 are included in the 4^th^-dose safety analysis set.

Demographic characteristics were comparable among treatment arms within adult PP-IMM (**Table 1**) and safety (**Table S1**) sets, other than a higher proportion of 18- to 64-year-old participants in the adult PP-IMM set who received a 3^rd^ dose, compared to other doses; characteristics were also comparable among treatment arms within the adolescent sets. Adult and adolescent participants received 3^rd^ or 4^th^ doses after a median of 6–12 months following the preceding dose (**Figure S1**).

**Table 1.** Demographic characteristics (PP-IMM analysis sets)

|  | Adult main study |  |  |  | Pediatric expansion (12 to <18 years) |  |  |  |
| --- | --- | --- | --- | --- | --- | --- | --- | --- |
|  | Primary series |  | 3<sup>rd</sup> Dose | 4<sup>th</sup> Dose | Primary series |  | 3<sup>rd</sup> Dose | 4<sup>th</sup> Dose |
| Characteristic | NVX-CoV2373<br>(n=713) | Placebo<br>(n=337) | NVX-CoV2373<br>(n=226) | NVX-CoV2373<br>(n=160) | NVX-CoV2373<br>(n=1126) | Placebo<br>(n=537) | NVX-CoV2373<br>(n=220) | NVX-CoV2373<br>(n=27) |
| **Sex, n (%)** |  |  |  |  |  |  |  |  |
| Male | 364 (51.1) | 162 (48.1) | 117 (51.8) | 76 (47.5) | 584 (51.9) | 297 (55.3) | 127 (57.7) | 16 (59.3) |
| Female | 349 (48.9) | 175 (51.9) | 109 (48.2) | 84 (52.5) | 542 (48.1) | 240 (44.7) | 93 (42.3) | 11 (40.7) |
| **Age (years)** |  |  |  |  |  |  |  |  |
| Mean (SD) | 57.1 (15.69) | 56.5 (16.81) | 49.6 (14.17) | 58.7 (13.89) | 13.8 (1.39) | 13.7 (1.42) | 14.1 (1.51) | 13.2 (1.22) |
| Median (range) | 64.0 (18–95) | 65.0 (18–85) | 52.0 (19–79) | 62.0 (19–84) | 14.0 (12–17) | 14.0 (12–17) | 14.0 (12–17) | 13.0 (12–17) |
| **Age group (years), n (%)** |  |  |  |  |  |  |  |  |
| 18–64 | 358 (50.2) | 168 (49.9) | 196 (86.7) | 91 (56.9) | N/A | N/A | N/A | N/A |
| ≥65 | 355 (49.8) | 169 (50.1) | 30 (13.3) | 69 (43.1) | N/A | N/A | N/A | N/A |
| 12 to <15 | N/A | N/A | N/A | N/A | 766 (68.0) | 368 (68.5) | 118 (53.6) | 23 (85.2) |
| 15 to <18 | N/A | N/A | N/A | N/A | 360 (32.0) | 169 (31.5) | 102 (46.4) | 4 (14.8) |
| **Region, n (%)** |  |  |  |  |  |  |  |  |
| Mexico | 48 (6.7) | 23 (6.8) | 0 | 0 | 0 | 0 | 0 | 0 |
| United States | 665 (93.3) | 314 (93.2) | 226 (100) | 160 (100) | 1126 (100) | 537 (100) | 220 (100) | 27 (100) |
| **Race, n (%)** |  |  |  |  |  |  |  |  |
| White | 571 (80.1) | 257 (76.3) | 187 (82.7) | 134 (83.8) | 866 (76.9) | 402 (74.9) | 186 (84.5) | 22 (81.5) |
| Black or African American | 62 (8.7) | 46 (13.6) | 21 (9.3) | 13 (8.1) | 142 (12.6) | 69 (12.8) | 12 (5.5) | 3 (11.1) |
| Asian | 19 (2.7) | 14 (4.2) | 13 (5.8) | 6 (3.8) | 34 (3.0) | 23 (4.3) | 7 (3.2) | 0 |
| American Indian or Alaska Native | 8 (6.7) | 16 (4.7) | 1 (0.4) | 5 (3.1) | 13 (1.2) | 6 (1.1) | 1 (0.5) | 0 |
| Mixed Origin | 10 (1.4) | 2 (0.6) | 3 (1.3) | 2 (1.3) | 63 (5.6) | 33 (6.1) | 12 (5.5) | 1 (3.7) |
| Native Hawaiian or Other Pacific Islander | 0 | 1 (0.3) | 1 (0.4) | 0 | 3 (0.3) | 1 (0.2) | 1 (0.5) | 0 |
| Not Reported | 3 (0.4) | 1 (0.3) | 0 | 0 | 5 (0.4) | 3 (0.6) | 1 (0.5) | 1 (3.7) |
| **Ethnicity, n (%)** |  |  |  |  |  |  |  |  |
| Hispanic or Latino | 142 (19.9) | 60 (17.8) | 34 (15.0) | 9 (5.6) | 175 (15.5) | 87 (16.2) | 36 (16.4) | 6 (22.2) |
| Not Hispanic or Latino | 570 (79.9) | 277 (82.2) | 192 (85.0) | 151 (94.4) | 947 (84.1) | 450 (83.8) | 184 (83.6) | 20 (74.1) |
| Not reported/unknown | 1 (0.1) | 0 | 0 | 0 | 4 (0.4) | 0 | 0 | 1 (3.7) |

SD, standard deviation. Note: Demographic and clinical characteristics are shown for the total PP-IMM set and were not developed for individual PP-IMM subsets.

### 3.2. Immunological responses to the ancestral strain in adults and adolescents

nAb (MN_50_) GMTs to the ancestral strain in adults rose from 11 to 1083 in the NVX-CoV2373 group (GMFR, 103 [95% CI, 92.4–115.6]) from day 0 to day 35 (**Figure 2A**). No antibody response was detected in placebo-recipients from a baseline GMT of 10. Day 35 SCRs were 94% with NVX-CoV2373 versus 2% with placebo. Additional doses of NVX-CoV2373 increased titers from pre-dose to 28 days post dose for both the 3^rd^ and 4^th^ doses, resulting in nAb titers greater than those observed 14 days post primary series. Similar response trends were observed across doses of NVX-CoV2373 in the adolescent population (**Figure 2B**). Noninferiority was achieved for the nAb response 28 days after the 3^rd^ dose compared to 14 days after the primary series (GMTR and SCR difference) in both the adult and adolescent populations (**Figure 2C**).

**Figure 2.**
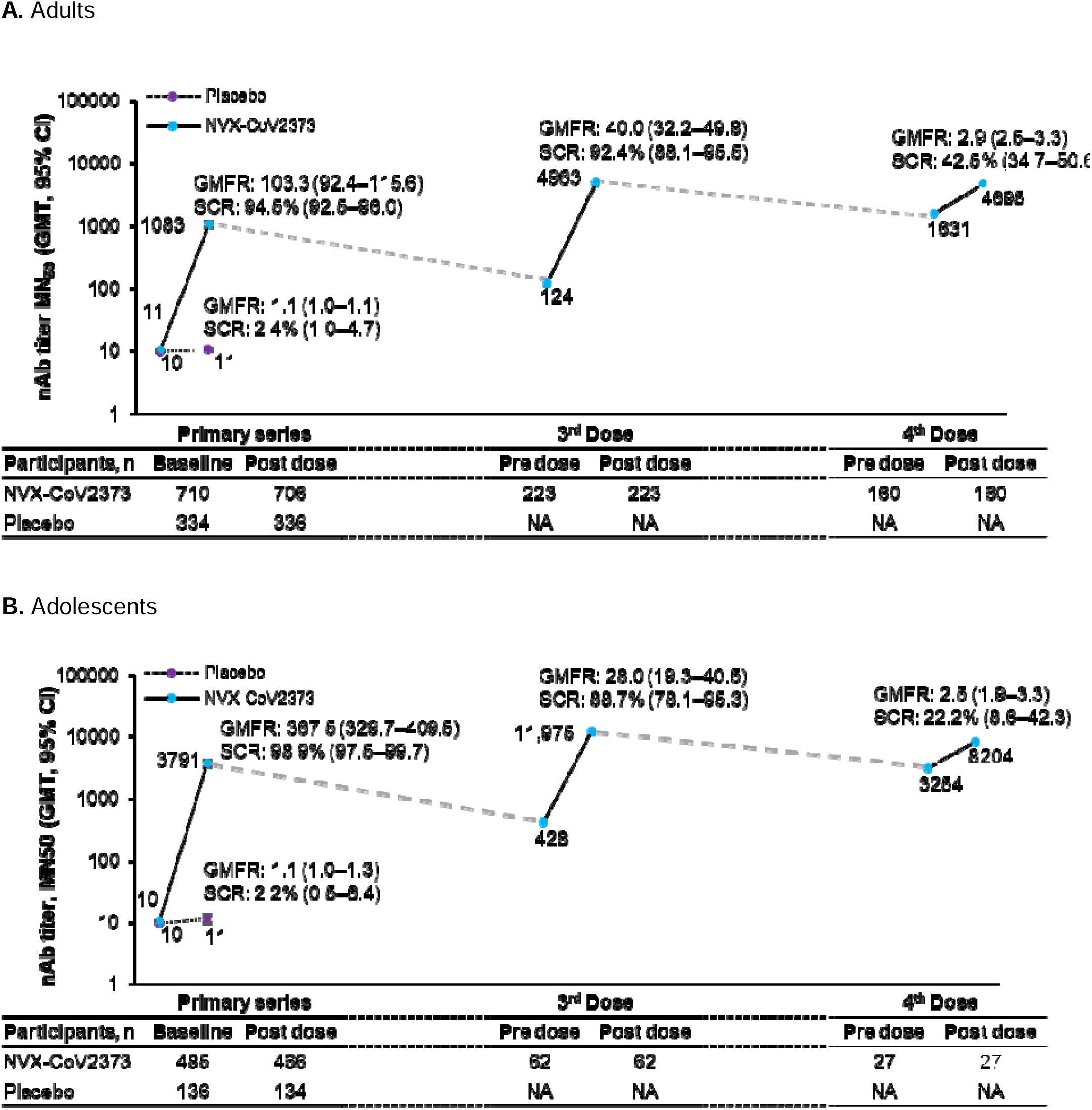
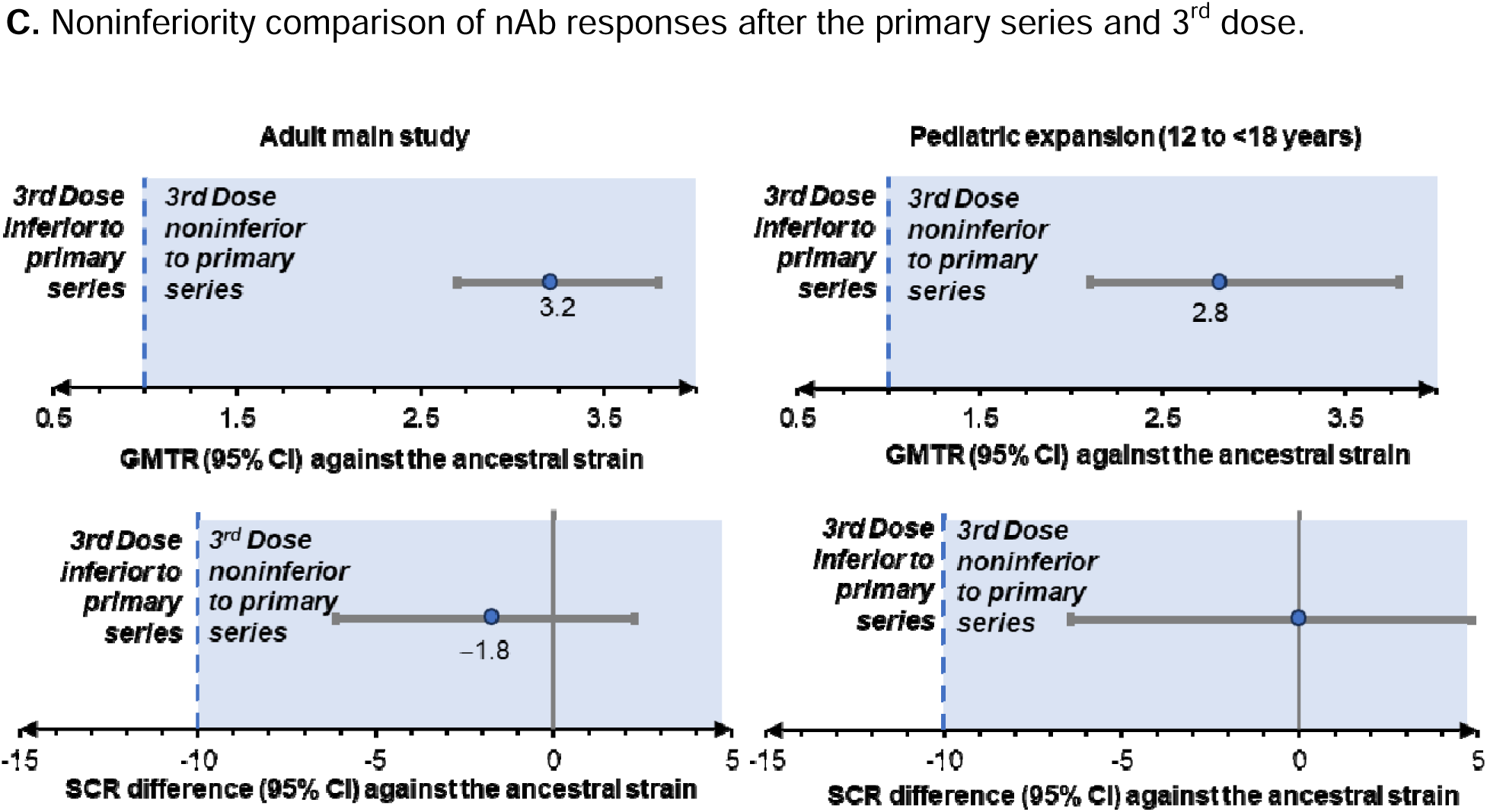
nAb responses against the ancestral strain in PP-IMM analysis sets. A) Adults. B) Adolescents. C) Noninferiority comparison of nAb responses after the primary series and 3rd dose. Baseline/pre-dose are defined as the last available assessment prior to administration of trial vaccine for the 1st, 3rd, or 4th dose. Post dose is 14 days (Day 35) after the primary series and 28 days after the 3rd or 4th dose. GMFR was calculated based on titer comparison of respective post vaccination and baseline samples for each group. SCR was calculated based on the number of participants with a _≥_4-fold increase in antibody concentration from baseline divided by the number of participants with a sample collected at the designated post vaccination time. GMFR and SCR values with 95% CIs are noted. To establish noninferiority, the following criteria were to be met: lower bound (LB) of two-sided 95% CI for the ratio of GMTs (GMTthird/GMTprimary) >1.0; and LB of the two-sided 95% CI for the difference of SCR (GMTthird − GMTprimary) > −10%. Responses are compared 14 days (Day 35) after the primary series and 28 days after the 3rd dose. Criteria boundaries for 95% CI are marked with a dashed vertical line. GMT, geometric mean titer; LB, lower bound; nAb, neutralizing antibody; SCR, seroconversion rate.

In adults, anti-rS IgG GMEUs rose from 119 to 49,384 in the NVX-CoV2373 group (GMFR, 414 [368.8–464.2]) from day 0 to 35 versus no apparent change from day 0 (GMEU, 108) in the placebo group (GMFR, 1.2 [95% CI, 1.1–1.3]) (**Figure S2A**). GMEUs also substantially increased from pre-dose to 28 days post 3^rd^ and 4^th^ doses. Adolescent participants had low baseline GMEUs on day 0 and a robust increase on day 35 post active vaccination (**Figure S2B**). Titers remained high with subsequent doses. hACE2 RBI GMTs followed a similar pattern as nAb and serum IgG responses in both adults (**Figure S3A**) and adolescents (**Figure S3B**). Immunological responses after the primary series among participants regardless of baseline serostatus were comparable to those in the baseline seronegative PP-IMM sets across each of the assays (**Figure S4**).

Durability of immunogenic response was assessed 4- and 8-months post 3^rd^ dose in subsets of the PP-IMM populations. Importantly, nAb, anti-rS IgG, and hACE2 RBI responses to the ancestral strain were preserved at levels comparable to those recorded 28 days after the 3^rd^ dose (**Figure S5**). Titers remained elevated from baseline and there were only minor declines in GMFR and SCRs through 8 months post 3^rd^ dose.

### 3.3. Immunological responses to the ancestral strain in adult age subgroups

Baseline nAb, serum IgG, and hACE2 RBI values were similar (within each assessment) among participants aged 18 to <65 years and ≥65 years in both treatment groups (**Table S2**). Fourteen days after the primary series, the nAb titers in the NVX-CoV2373 group were ∼1.4-fold higher in participants aged 18 to <65 years (123.8 [107.1–143.1]) compared with those aged ≥65 years (86.1 [72.6–102.1]). Increases in serum IgG and hACE2 RBI titers on day 35 followed a similar trend. SCRs from day 0 to day 35 for each of the response measures were comparable among the adult age groups. Immune responses in adults who received a 3^rd^ and/or 4^th^ dose of NVX-CoV2373 were similar in younger and older adults (**Table S3**).

### 3.4. Cross-reactivity of NVX-CoV2373 against Omicron subvariants

nAb responses (ID_50_) to BA.5 and BQ.1.1 were assessed in a PP-IMM subset of adult participants who received all four study doses of NVX-CoV2373. Fourteen days after the primary series, responses that cross-reacted with variant strains were ∼100-fold lower than those against the ancestral strain; however, activity increased against each variant with subsequent doses of NVX-CoV2373 (**Figure 3A**).

**Figure 3.**
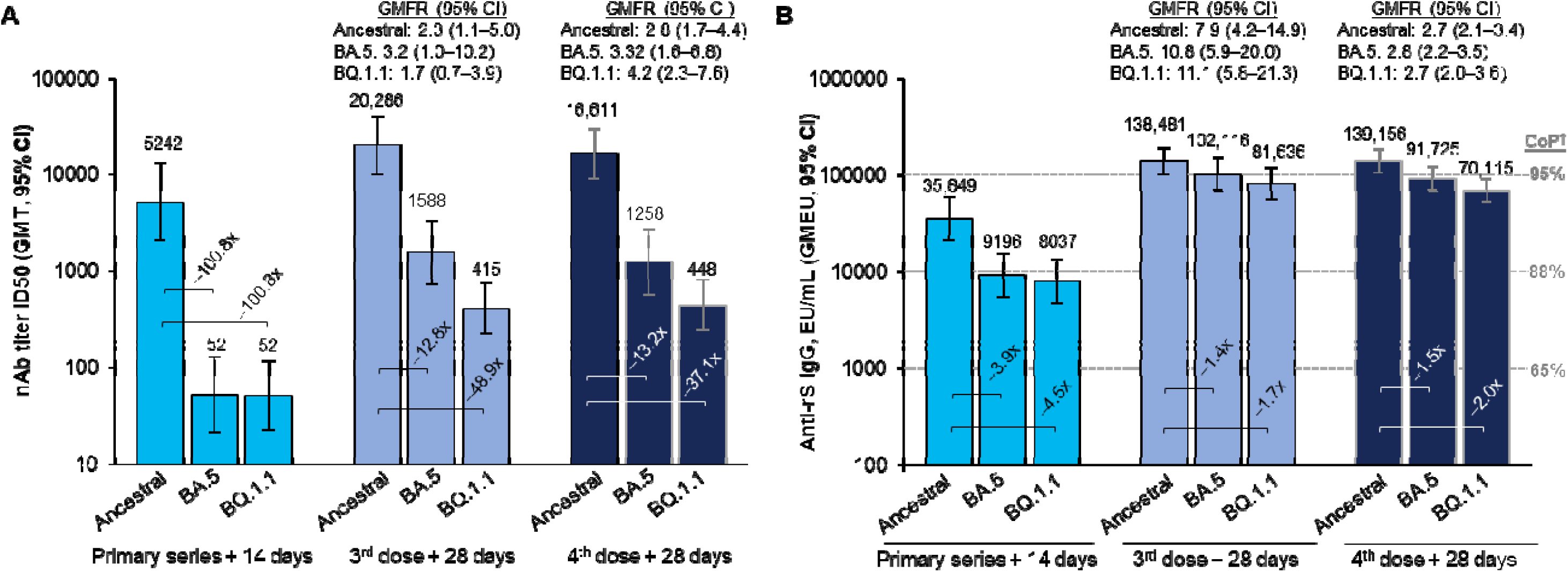
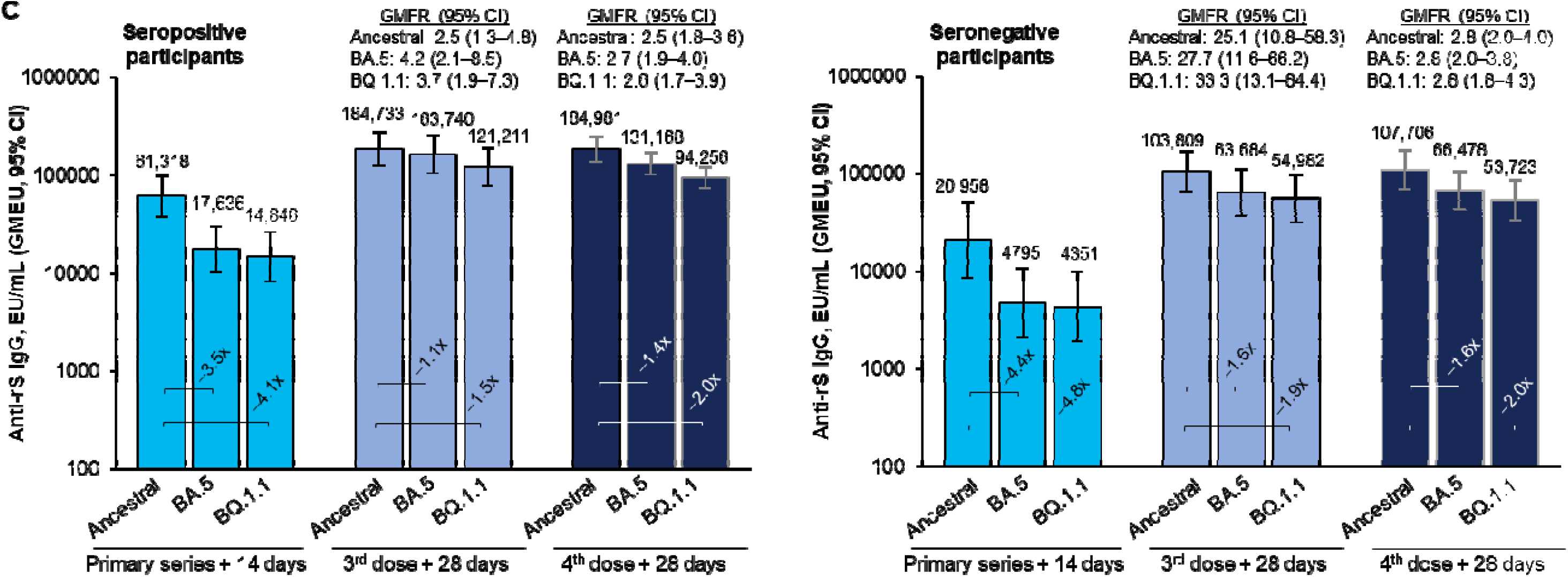
Immunogenicity in an adult PP-IMM subset against the ancestral strain, BA.5, and BQ.1.1 before and after the primary series, 3^rd^ dose, and 4^th^ dose. A) Pseudovirus neutralization assay (n=15 participants for each timepoint and strain). B) Anti-rS IgG responses regardless of baseline SARS-CoV-2 serostatus (n=40 participants for each timepoint and strain, other than for 38 participants for 28 days after the 4^th^ dose). C) Anti-rS IgG responses by baseline SARS-CoV-2 serostatus (n=20 participants for each timepoint and strain, other than for 18 participants for 28 days after the 4^th^ dose). Fold differences compared with the ancestral strain for each series are shown in brackets. GMFR are comparing GMTs from samples collected on the day of dosing (before vaccination) with samples collected 28 days after that same dose. CoP values in (B) were determined for the ancestral strain based on Day 35 anti-rS IgG levels with protection 56 days later (Fong et al 2022). CoP, correlate of protection; GMFR, geometric mean fold rise; GMT, geometric mean titer; SCR, seroconversion rate.

Cross-reactive serum anti-rS IgG responses were observed in adults against BA.5 and BQ.1.1 after the primary series (∼4-fold lower than against the ancestral virus) and subsequent doses (∼1.5-fold lower than those against ancestral virus; **Figure 3B**). Anti-rS IgG levels against the ancestral strain and variants aligned with correlates of protection of >88% (based on day 35 anti-rS IgG levels and protection 56 days later), as described previously.^16^ Anti-rS IgG responses were also assessed based on positive or negative SARS-CoV-2 NP serostatus at baseline, and a similar trend was observed as those described for the full immunogenicity subset (**Figure 3C**).

### 3.5. Reactogenicity

Overall, solicited AEs were predominantly mild to moderate in severity and self-limited. Events occurred at a somewhat higher frequency after the 3^rd^ and 4^th^ doses compared with the first two doses (**Figure 4A; Table S4**). Solicited AEs were reported most frequently within 2 days after vaccination and lasted a median 1–3 days; there were a limited number of AEs lasting >7 days post injection (**Table S5**). Frequencies of solicited local AEs across doses trended similarly in both the adult and adolescent study populations; following each dose, pain and tenderness were the most common (>10%). Any solicited systemic AEs also occurred with similar frequency among the adult and adolescent study populations (**Figure 4B; Table S4**). Fatigue, headache, malaise, and muscle pain were the most common (>40% for any dose) solicited systemic AEs reported.

**Figure 4.**
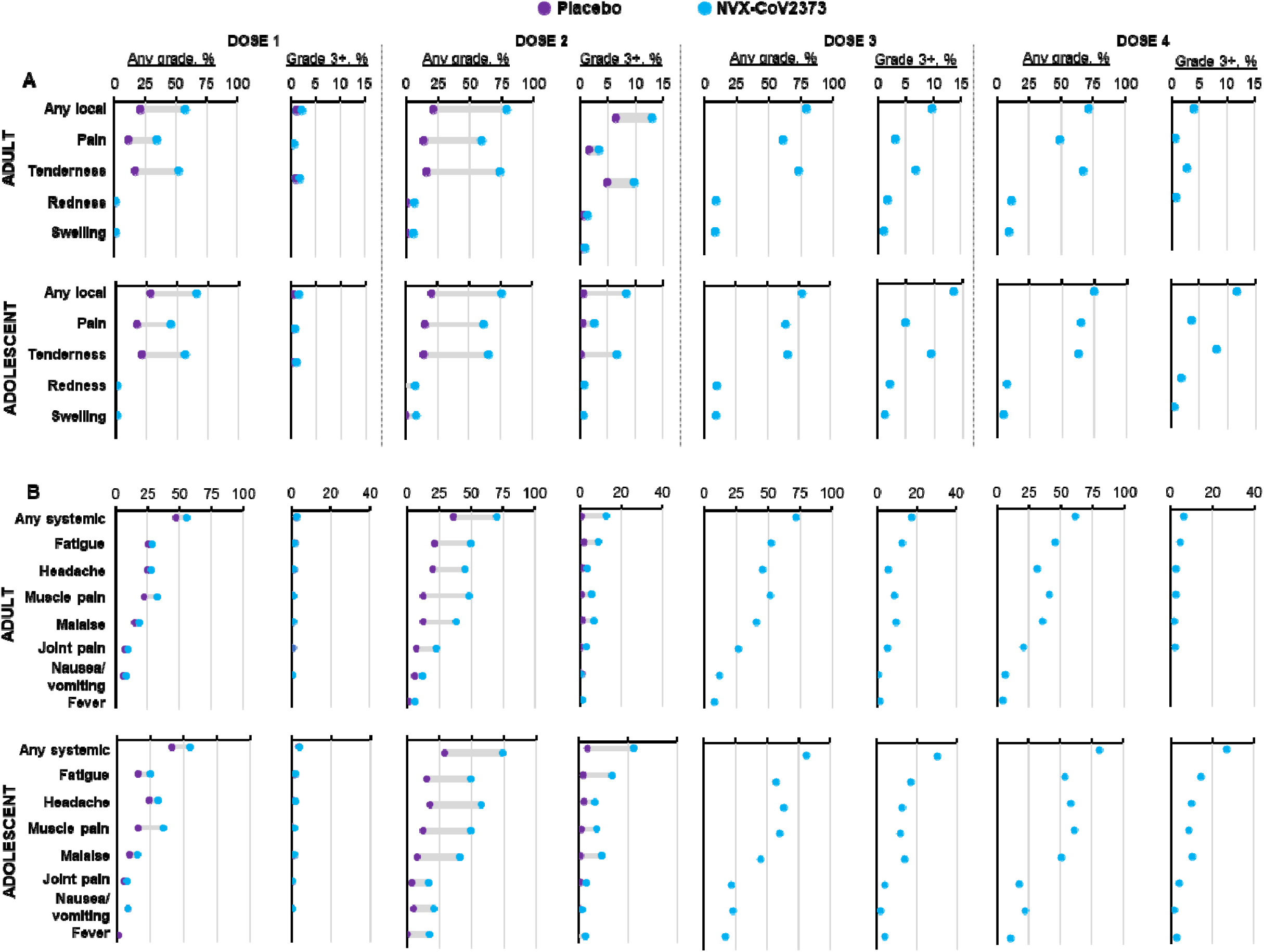
Reactogenicity within 7 days of each study dose. Solicited (A) local and (B) systemic treatment-emergent adverse events were collected in adult (top) and adolescent (bottom) participants in the safety analysis sets with at least one eDiary entry. Note: There are no placebo reactogenicity data beyond the primary series as subsequent doses of placebo were not administered.

### 3.6. Unsolicited AEs

Through 28 days after the primary series, unsolicited AEs occurred at similar frequencies within the adult and adolescent populations in the NVX-CoV2373 (12.1% and 15.8%, respectively) and placebo (11.6% and 16.0%) groups (**Table S6**). Frequency of unsolicited AEs decreased with the 3^rd^ and 4th doses of NVX-CoV2373 in both adults and adolescents. After the 4^th^ dose, no adults and one (0.5%) adolescent participant had a vaccine-related AE (urticaria). Overall, there was minimal occurrence of vaccine-related or severe AEs, as well as MAAEs, SAEs, and PIMMCs. During primary series follow-up, six adult participants discontinued the study for an unsolicited AE considered related to study vaccination. After unblinding, it was determined these were four and two participants in the active vaccine and placebo groups, respectively. At the data cutoff date, there were no unsolicited AEs leading to discontinuation in adolescent participants. No deaths during the study were considered related to vaccination, and there were no deaths in the adolescent population.

## 4. Discussion

These results expand on previous reports of the PREVENT-19 trial^8,10^ and support phase 3 data^17^ that demonstrated the safety and efficacy of two primary doses of NVX-CoV2373 in adults and adolescents. Immunological responses to ancestral SARS-CoV-2 were markedly increased 14 days after two primary doses of NVX-CoV2373 in participants serologically negative for SARS-CoV-2 NP. Responses were durable, with continued robust humoral responses after a 3^rd^ dose of NVX-CoV2373 and modest increases after a 4^th^ dose. Over the full period of the study, antibody titers were sustained in the range associated with a high degree of protective efficacy.^16^ Overall, NVX-CoV2373 was immunogenic in both younger and older adults and adolescents and was well tolerated.

In both adults and adolescents, immunological response to a 3^rd^ dose of NVX-CoV2373 was noninferior to that observed 14 days after the primary series. A median of 8 months after the 3^rd^ dose, a 4^th^ dose of NVX-CoV2373 resulted in elevated nAb titers comparable to those post 3^rd^ dose, and >4-fold higher than those observed 28-days post primary series. These positive 3^rd^-dose responses are in line with those previously reported from a phase 2 trial.^18^ nAb titers before the 3^rd^ and 4^th^ doses were higher than those reported on Day 0, indicating response durability. Notably, there was some decline in titers, indicative of waning immunity; however, this can be expected as it would align with previously reported waning vaccine efficacy (to Delta variants) >120 days post vaccination.^19^ Although mRNA COVID-19 vaccines have generally reported higher absolute post-vaccination IgG levels and neutralization titers, other studies have demonstrated significantly better persistence of response with NVX-CoV2373, with a slower decline in immune responses.^20^ These data support the rationale for periodic reimmunization to help combat breakthrough infection in vaccinated individuals, especially with the frequent emergence of new variant strains.

Overall, AEs were mild to moderate and reactogenicity was generally transient. Incidence and severity of local and systemic reactogenicity moderately increased following the 2^nd^ dose and remained relatively consistent after both the 3^rd^ dose and 4^th^ dose. These findings align with early studies during the pandemic where a COVID-19 vaccine primary series (ie, mRNA, adenoviral vector, and protein-based) was observed to generally elicit higher frequencies of reactogenic symptoms following the 2^nd^ dose compared to the 1^st^ dose.^11,21,22^ Side effects from COVID-19 vaccines have been identified as a driving force of vaccine hesitancy;^23,24^ therefore, a key observation from this study may be that additional doses of NVX-CoV2373 in adult and adolescent populations did not further increase reactogenic symptom frequency. Here, unsolicited AE frequency after the primary series was comparable in the NVX-CoV2373 and placebo groups and incidence appeared to plateau after the 4^th^ dose, which is consistent with data from a phase 2 study conducted in Australia and the US.^21^ Notably, the solicited and unsolicited AE outcomes after repeat dosing with NVX-CoV2373 are reassuring for future recommendations of additional doses of the Novavax COVID-19 vaccine, based on updated formulations that are expected due to continued SARS-CoV-2 strain circulation and evolution.

A tolerable safety profile, immunogenicity, and protection from infection in older adults is of great interest for COVID-19 vaccines due to the higher risk of severe disease complications in those aged ≥65 years.^25^ No safety concerns were raised, and high and similar immune responses were observed across age groups; however, as expected, titers were lower in the older versus younger adult population, and titers were higher for the adolescent versus adult populations. Notably, in both age groups, responses were within the range demonstrated to provide protective efficacy.^16^ This is an important consideration, as real-world evidence indicates that vaccine side effects are a key factor in selection of a specific vaccine formulation when efficacy among the options is comparable.^26^ Overall, the decision for use of a protein- or mRNA-based vaccine formulation should consider patient characteristics/comorbidities and individual preference (eg, vaccine hesitancy due to side effects), as well as immunogenicity.

Cross-protective humoral immune responses were observed against Omicron variants BA.5 and BQ.1.1. Over time, SARS-CoV-2 lineages have evolved, attaining increased fitness and immune evasion capabilities, compared to the ancestral strain and early Omicron variants. For example, additional mutations in the viral spike protein may promote improved replication in respiratory cells, decreased affinity for ACE2, and antibody resistance.^27^ A 3^rd^ dose of NVX-CoV2373 elicited a robust serum IgG response to BA.1 with an SCR comparable to that observed after the primary series. These data are in line with those from a phase 2 study of a smaller South African population, where elevated titers against BA.1 and BA.4/5 were observed after 3^rd^ and 4^th^ doses of NVX-CoV2373.^28^ These immunogenicity results provide supportive evidence for this protein-based vaccine platform’s capacity to elicit broadly cross-reactive nAb titers against antigenically distant SARS-CoV-2 variant strains.

This study was conducted during the early years of what has become a continuously evolving epidemiology of SARS-CoV-2. At study initiation, virtually all participants were infection-naive, and ancestral virus, as well as some early antigenically related variants, were circulating. During the second year, more antigenically divergent variants emerged (eg, BA.1, BA.5, and BQ.1.1), reducing immunity among individuals vaccinated with authorized prototype vaccines. This led to recommendations for “booster” doses of the prototype vaccine, followed by updated vaccine formulations that targeted newer variants. For the 2023–2024 season, vaccines based on the XBB.1.5 variant became available.^2,3^ JN.1, followed by strains within that lineage (ie, KP.2 and KP.3), then became the dominant strains, and two mRNA-based vaccines and the Novavax COVID-19 vaccine were authorized/approved as updated COVID-19 vaccines for the 2024–2025 season.^7^ In non-clinical models, the updated Novavax COVID-19 vaccine (NVX-CoV2705 [JN.1]) generated broad cross-neutralizing responses against JN.1-lineage subvariants, including KP.2 and KP.3.^29^ The continued authorizations, preclinical data, and assessments with convalescent serum samples suggest consistency for immunogenicity and effectiveness of updated Novavax COVID-19 vaccines with this platform.

Immune responses described here demonstrate cross-reactivity of the prototype vaccine with genetic variants that had deviated from ancestral SARS-CoV-2. It can be expected that immunogenicity of prototype vaccines would decline as new strains emerge and antigenic distance grows. For example, based on genomic composition, early Omicron strains (eg, BA.1, BA.5, and BQ.1) may be considered one antigenically related “family”, followed by a larger shift to XBB-related strains that are more antigenically distinct from ancestral SARS-CoV-2.^30^ Notably, within these ‘antigenic families’ of variants, cross-reactivity to updated vaccines may be more likely.

A separate analysis of data from PREVENT-19 reported that nAb titers provide a correlate of protection for NVX-CoV2373 against ancestral virus,^16^ and a more recent analysis^19^ confirmed antibody levels to the immune-evading Delta variant that support protective efficacy. This vaccine platform also demonstrated cross-reactivity in the 2019nCoV-313/NCT05975060 study investigating an updated version of NVX-CoV2373 directed to XBB.1.5 (NVX-CoV2601), [*in press*], which was authorized for use in 2023–2024.^2,3^ In that study, nAb responses to JN.1 and KP.2 were observed in previously vaccinated participants (≥3 doses of an mRNA-based COVID-19 vaccine) who received one dose of NVX-CoV2601. Cross-reactivity against SARS-CoV-2 variants by vaccines generated using this platform may be related to unique platform attributes, such as the saponin-based Matrix-M adjuvant and/or greater accessibility to highly conserved spike protein epitopes due to the truncated glycans of glycoproteins produced in the baculovirus expression vector system.^31^ Matrix-M adjuvant likely broadens epitope recognition^32^ and elicits a Th1-dominant immune response capable of improving humoral immune responses^33^. Despite the continued evolution of SARS-CoV-2, the ancestral vaccine and strain responses analyzed in this study allow for the interpretation of cross-reactivity specific to this vaccine platform, which can provide insight into potential performance of updated vaccine formulations against antigenically distant future-drifted strains.

## 5. Conclusions

In conclusion, NVX-CoV2373 elicited robust and durable humoral immune responses to SARS-CoV-2, as well as cross-protective responses to key variants. Doses of NVX-CoV2373 beyond the primary series induced spike-specific antibodies, with an acceptable reactogenicity profile. These results support continuing updates to this vaccine platform based on currently circulating strains to help effectively combat SARS-CoV-2 infection.

## Data Availability

Study details can be found at https://clinicaltrials.gov/study/NCT04611802. Data that support the findings of this study are available in supplementary material and from the corresponding author upon reasonable request.

## Abbreviations

AE: adverse event
AESI: adverse event of special interest;
ELISA: enzyme-linked immunosorbent assay;
hACE2-RBI: human angiotensin-converting enzyme-2 receptor binding inhibition;
GMEU: geometric mean ELISA unit;
GMFR: geometric mean fold rise;
GMT: geometric mean titer;
LLOQ: lower limit of quantification;
MAAE: medically attended adverse event;
nAb: neutralizing antibody;
NP: nucleoprotein;
PIMMC: potentially immune-mediated medical condition;
PP-IMM: per-protocol immunogenicity;
rS: recombinant Spike;
SAE: serious adverse event;
SCR: seroconversion rate;
TEAE: treatment-emergent adverse event.

## Acknowledgments

We especially thank each of the study participants who volunteered for this study, as well as the 2019nCoV-301 Study Group members.^8,10^ We acknowledge the funders (BARDA and NIAID/NIH), the members of the NIAID Data and Safety Monitoring Board (DSMB), community leadership groups throughout the country who assisted with community engagement and recruitment, unnamed colleagues at each of the sites who generously contributed to the trials in many ways, and all unnamed colleagues at Novavax, Inc, who worked tirelessly and gave unlimited efforts to the development, testing, and support of this trial. Medical writing support was provided by Kelly Cameron, PhD, CMPP, Kelly M. Fahrbach, PhD, CMPP, with editorial support from Ebenezer M. Awuah-Yeboah, BS, all of Ashfield MedComms (New York, USA), an Inizio company, supported by Novavax, Inc.

## Appendix A

Supplementary data

Supplementary study results.

## Ethics Approval Statement

The study was performed in accordance with the International Conference on Harmonization Good Clinical Practice guidelines and overseen by the NIAID Data Safety and Monitoring Board.

## Patient Consent Statement

All participants provided written informed consent before enrollment.

## Author contributions

**Germán Áñez**: conceptualization, methodology, formal analysis, investigation, writing – original draft preparation, writing – reviewing and editing

**Alice McGarry**: formal analysis, investigation, writing – reviewing and editing

**Wayne Woo**: formal analysis, investigation, writing – reviewing and editing

**Karen L. Kotloff**: data curation, investigation, writing – reviewing and editing; resources

**Cynthia L. Gay**: data curation, investigation, writing – reviewing and editing; resources

**Mingzhu Zhu**: writing – reviewing and editing; resources

**Shane Cloney-Clark**: formal analysis, writing – reviewing and editing

**Joy Nelson**: writing – reviewing and editing; project administration

**Haoua Dunbar**: data curation, writing – reviewing and editing

**Miranda R. Cai**: formal analysis, writing – reviewing and editing

**Iksung Cho**: formal analysis, investigation, writing – reviewing and editing, software

**Zhaohui Cai**: formal analysis, writing – reviewing and editing

**Raj Kalkeri**: investigation, writing – reviewing and editing

**Joyce S. Plested**: formal analysis, writing – reviewing and editing

**Nita Patel**: resources, writing – reviewing and editing

**Katherine Smith**: formal analysis, writing – reviewing and editing

**Anthony M. Marchese**: writing – updated draft preparation, writing – reviewing and editing

**Gregory M. Glenn**: methodology, investigation, writing – reviewing and editing

**Raburn M. Mallory**: methodology, formal analysis, writing – reviewing and editing

**Lisa M. Dunkle**: conceptualization, methodology, formal analysis, investigation, writing – reviewing and editing, writing – original draft preparation

## References

1. COVID-19 Vaccine Tracker. Novavax: Nuvaxovid https://covid19.trackvaccines.org/vaccines/25/. Accessed January 16, 2024.

2. Food and Drug Administration. Novavax COVID-19 Vaccine, Adjuvanted. Available online 17 October 2023. https://www.fda.gov/vaccines-blood-biologics/coronavirus-covid-19-cber-regulated-biologics/novavax-covid-19-vaccine-adjuvanted. Accessed January 31, 2024.

3. European Medicines Agency. Nuvaxovid. COVID-19 Vaccine (recombinant, adjuvanted). https://www.ema.europa.eu/en/medicines/human/EPAR/nuvaxovid#ema-inpage-item-product-info. Accessed February 1, 2024.

4. World Health Organization. Updated Risk Evaluation of JN.1, 15 April 2024. https://www.who.int/docs/default-source/coronaviruse/15042024_jn1_ure.pdf?sfvrsn=8bd19a5c_7#:~:text=Effectiveness%20of%20the%20XBB.,predicted%20to%20cross%2Drecognize%20BA. Accessed May 13, 2024.

5. Link-Gelles R, Ciesla AA, Mak J, et al. Early Estimates of Updated 2023-2024 (Monovalent XBB.1.5) COVID-19 Vaccine Effectiveness Against Symptomatic SARS-CoV-2 Infection Attributable to Co-Circulating Omicron Variants Among Immunocompetent Adults - Increasing Community Access to Testing Program, United States, September 2023-January 2024. MMWR Morb Mortal Wkly Rep. 2024;73(4):77–83.

6. U.S. Food and Drug Administration. Updated COVID-19 Vaccines for Use in the United States Beginning in Fall 2024 https://www.fda.gov/vaccines-blood-biologics/updated-covid-19-vaccines-use-united-states-beginning-fall-2024. Accessed September 13, 2024.

7. U.S. Food and Drug Administration. COVID-19 Vaccines. https://www.fda.gov/emergency-preparedness-and-response/coronavirus-disease-2019-covid-19/covid-19-vaccines. Accessed September 25, 2024.

8. Dunkle LM, Kotloff KL, Gay CL, et al. Efficacy and Safety of NVX-CoV2373 in Adults in the United States and Mexico. N Engl J Med. 2022;386(6):531-543.

9. Marchese AM, Zhou X, Kinol J, et al. NVX-CoV2373 vaccine efficacy against hospitalization: A post hoc analysis of the PREVENT-19 phase 3, randomized, placebo-controlled trial. Vaccine. 2023;41(22):3461–3466.

10. Anez G, Dunkle LM, Gay CL, et al. Safety, Immunogenicity, and Efficacy of the NVX-CoV2373 COVID-19 Vaccine in Adolescents: A Randomized Clinical Trial. JAMA Netw Open. 2023;6(4):e239135.

11. Baden LR, El Sahly HM, Essink B, et al. Efficacy and Safety of the mRNA-1273 SARS-CoV-2 Vaccine. N Engl J Med. 2021;384(5):403–416.

12. Hamilton S, Zhu M, Cloney-Clark S, et al. Validation of a severe acute respiratory syndrome coronavirus 2 microneutralization assay for evaluation of vaccine immunogenicity. J Virol Methods. 2024;327:114945.

13. Zhu M, Cloney-Clark S, Feng SL, et al. A Severe Acute Respiratory Syndrome Coronavirus 2 Anti-Spike Immunoglobulin G Assay: A Robust Method for Evaluation of Vaccine Immunogenicity Using an Established Correlate of Protection. Microorganisms. 2023;11(7).

14. Plested JS, Zhu M, Cloney-Clark S, et al. Severe Acute Respiratory Syndrome Coronavirus 2 Receptor (Human Angiotensin-Converting Enzyme 2) Binding Inhibition Assay: A Rapid, High-Throughput Assay Useful for Vaccine Immunogenicity Evaluation. Microorganisms. 2023;11(2).

15. Cai Z, Kalkeri R, Zhu M, et al. A Pseudovirus-Based Neutralization Assay for SARS-CoV-2 Variants: A Rapid, Cost-Effective, BSL-2-Based High-Throughput Assay Useful for Vaccine Immunogenicity Evaluation. Microorganisms. 2024;12(3).

16. Fong Y, Huang Y, Benkeser D, et al. Immune correlates analysis of the PREVENT-19 COVID-19 vaccine efficacy clinical trial. Nat Commun. 2023;14(1):331.

17. Heath PT, Galiza EP, Baxter DN, et al. Safety and Efficacy of the NVX-CoV2373 Coronavirus Disease 2019 Vaccine at Completion of the Placebo-Controlled Phase of a Randomized Controlled Trial. Clin Infect Dis. 2023;76(3):398–407.

18. Mallory RM, Formica N, Pfeiffer S, et al. Safety and immunogenicity following a homologous booster dose of a SARS-CoV-2 recombinant spike protein vaccine (NVX-CoV2373): a secondary analysis of a randomised, placebo-controlled, phase 2 trial. Lancet Infect Dis. 2022;22(11):1565–1576.

19. Follmann D, Mateja A, Fay MP, et al. Durability of Protection Against COVID-19 Through the Delta Surge for the NVX-CoV2373 Vaccine. Clin Infect Dis. 2024;79(1):78–85.

20. Liu X, Munro APS, Wright A, et al. Persistence of immune responses after heterologous and homologous third COVID-19 vaccine dose schedules in the UK: eight-month analyses of the COV-BOOST trial. J Infect. 2023;87(1):18–26.

21. Alves K, Plested JS, Galbiati S, et al. Immunogenicity and safety of a fourth homologous dose of NVX-CoV2373. Vaccine. 2023;41(29):4280–4286.

22. Polack FP, Thomas SJ, Kitchin N, et al. Safety and Efficacy of the BNT162b2 mRNA Covid-19 Vaccine. N Engl J Med. 2020;383(27):2603–2615.

23. Al-Obaydi S, Hennrikus E, Mohammad N, Lehman EB, Thakur A, Al-Shaikhly T. Hesitancy and reactogenicity to mRNA-based COVID-19 vaccines-Early experience with vaccine rollout in a multi-site healthcare system. PLoS One. 2022;17(8):e0272691.

24. Chrissian AA, Oyoyo UE, Patel P, et al. Impact of COVID-19 vaccine-associated side effects on health care worker absenteeism and future booster vaccination. Vaccine. 2022;40(23):3174–3181.

25. Centers for Disease Control and Prevention. People With Certain Medical Conditions. https://www.cdc.gov/coronavirus/2019-ncov/need-extra-precautions/people-with-medical-conditions.html. Accessed January 31, 2024.

26. Salisbury D, Lazarus JV, Waite N, et al. COVID-19 Vaccine Preferences in General Populations in Canada, Germany, the United Kingdom, and the United States: Discrete Choice Experiment. JMIR Public Health Surveill. 2024;10:e57242.

27. Planas D, Staropoli I, Michel V, et al. Distinct evolution of SARS-CoV-2 Omicron XBB and BA.2.86/JN.1 lineages combining increased fitness and antibody evasion. Nat Commun. 2024;15(1):2254.

28. Bhiman JN, Richardson SI, Lambson BE, et al. Novavax NVX-COV2373 triggers neutralization of Omicron sub-lineages. Sci Rep. 2023;13(1):1222.

29. Walker R. Vaccines and Related Biological Products Advisory Committee. Novavax Data in Support of 2024-2025 Vaccine Update. June 5, 2024. Available at: https://www.fda.gov/media/179143/download. Accessed October 20, 2024.

30. Nextstrain.org. Genomic epidemiology of SARS-CoV-2 with subsampling focused globally since pandemic start. https://nextstrain.org/ncov/gisaid/global/all-time?m=div. Accessed August 12, 2024.

31. Wolff MW, Murhammer DW, Jarvis DL, Linhardt RJ. Electrophoretic analysis of glycoprotein glycans produced by lepidopteran insect cells infected with an immediate early recombinant baculovirus encoding mammalian beta1,4-galactosyltransferase. Glycoconj J. 1999;16(12):753–756.

32. Chung KY, Coyle EM, Jani D, et al. ISCOMATRIX adjuvant promotes epitope spreading and antibody affinity maturation of influenza A H7N9 virus like particle vaccine that correlate with virus neutralization in humans. Vaccine. 2015;33(32):3953–3962.

33. Stertman L, Palm AE, Zarnegar B, et al. The Matrix-M adjuvant: A critical component of vaccines for the 21(st) century. Human vaccines & immunotherapeutics. 2023;19(1):2189885.

