## Supplementary material for "Safety and immunogenicity of four sequential doses of NVX-CoV2373 in adults and adolescents: a phase 3, randomized, placebo-controlled trial (PREVENT-19)": N/A

**Table S1.** Demographic characteristics (safety analysis sets).

|  | Adult main study | | | | | | Pediatric expansion (12 to <18 years) | | | |
| --- | --- | --- | --- | --- | --- | --- | --- | --- | --- | --- |
|  | Primary series | | 3^rd^ dose | | 4^th^ dose | | Primary series | | 3rd dose | 4^th^ dose |
| Parameter | NVX-CoV2373  (n=19,735) | Placebo  (n=9847) | | NVX-CoV2373  (n=13,354) | **NVX-CoV2373**  (n=359) | | **NVX-CoV2373**  (n=1487) | **Placebo**  (n=745) | NVX-CoV2373 (n=1499) | NVX-CoV2373 (n=205) |
| **Age (years)** | | | | | | | | | | |
| Mean (SD) | 46.5 (15.05) | 46.8 (14.95) | | 48.7 (14.76) | 58.7 (13.06) | | 13.9 (1.41) | 13.8 (1.44) | 13.8 (1.41) | 13.7 (1.32) |
| Median (range) | 47.0 (18–95) | 47.0 (18–90) | | 50.0 (18–96) | 61.0 (20–85) | | 14.0 (12–17) | 14.0 (12–17) | 14.0 (12–17) | 13.0 (12–17) |
| **Age group, n (%)** | | | | | | | | | | |
| 18 to <65 years | 17,255 (87.4) | 8612 (87.5) | | 11,290 (84.5) | 224 (62.4) | | N/A | N/A | N/A | N/A |
| ≥65 years | 2480 (12.6) | 1235 (12.5) | | 2064 (15.5) | 135 (37.6) | | N/A | N/A | N/A | N/A |
| 12 to <15 | N/A | N/A | | N/A | N/A | | 997 (67.0) | 500 (67.1) | 1,020 (68.0) | 146 (71.2) |
| 15 to <18 | N/A | N/A | | N/A | N/A | | 490 (33.0) | 245 (32.9) | 479 (32.0) | 59 (28.8) |
| **Sex, n (%)** | | | | | | | | | | |
| Male | 10,367 (52.5) | 5019 (51.0) | | 6764 (50.7) | 172 (47.9) | | 756 (50.8) | 416 (55.8) | 806 (53.8) | 102 (49.8) |
| Female | 9368 (47.5) | 4828 (49.0) | | 6590 (49.3) | 187 (52.1) | | 731 (49.2) | 329 (44.2) | 693 (46.2) | 103 (50.2) |
| **Race, n (%)** | | | | | | | | | | |
| White | 14,794 (75.0) | 7,381 (75.0) | | 9585 (71.8) | 285 (79.4) | | 1,115 (75.0) | 545 (73.2) | 1,096 (73.1) | 152 (74.1) |
| Black or African American | 2320 (11.8) | 1,164 (11.8) | | 1887 (14.1) | 50 (13.9) | | 202 (13.6) | 108 (14.5) | 219 (14.6) | 31 (15.1) |
| American Indian or Alaskan Native | 1309 (6.6) | 660 (6.7) | | 1044 (7.8) | 7 (1.9) | | 32 (2.2) | 14 (1.9) | 40 (2.7) | 0 |
| Asian | 809 (4.1) | 416 (4.2) | | 496 (3.7) | 12 (3.3) | | 43 (2.9) | 34 (4.6) | 53 (3.5) | 9 (4.4) |
| Mixed origin | 329 (1.7) | 160 (1.6) | | 225 (1.7) | 4 (1.1) | | 82 (5.5) | 37 (5.0) | 77 (5.1) | 10 (4.9) |
| Native Hawaiian/Other Pacific Islander | 56 (0.3) | 12 (0.1) | | 30 (0.2) | 0 | | 3 (0.2) | 2 (0.3) | 5 (0.3) | 1 (0.5) |
| Not reported | 110 (0.6) | 47 (0.5) | | 83 (0.6) | 1 (0.3) | | 10 (0.7) | 5 (0.7) | 9 (0.6) | 2 (1.0) |
| Missing | 8 (< 0.1) | 7 (< 0.1) | | 4 (<0.1) | 0 | | 0 | 0 | 0 | 0 |
| **Ethnicity, n (%)** | | | | | | | | | | |
| Hispanic or Latino | 4334 (22.0) | 2155 (21.9) | | 3,121 (23.4) | 22 (6.1) | 274 (18.4) | | 138 (18.5) | 276 (18.4) | 33 (16.1) |
| Not Hispanic or Latino | 15,345 (77.8) | 7669 (77.9) | | 10,196 (76.4) | 336 (93.6) | 1,208 (81.2) | | 607 (81.5) | 1,220 (81.4) | 171 (83.4) |
| Not reported | 32 (0.2) | 19 (0.2) | | 22 (0.2) | 1 (0.3) | 2 (0.1) | | 0 | 1(<0.1) | 0 |
| Unknown | 22 (0.1) | 3 (<0.1) | | 14 (0.1) | 0 | 3 (0.2) | | 0 | 2 (0.1) | 1 (0.5) |
| Missing | 2 (<0.1) | 1 (<0.1) | | 1 (<0.1) | 0 | 0 | | 0 | 0 | 0 |

Characteristics are based on populations who received NVX-CoV2373 in the initial vaccination period (pre crossover).

N/A, not applicable; SD, standard deviation.

**Table S2.** Immunological responses to the ancestral strain before and after primary series immunizations in age subgroups (PP-IMM analysis sets).

|  | **Aged 18 to <65 years** | | **Aged ≥65 years** | |
| --- | --- | --- | --- | --- |
|  | **NVX-CoV2373 (n=358)** | **Placebo (n=168** | **NVX-CoV2373**  **(n=355)** | **Placebo**  **(n=169)** |
| **nAb (MN_50_) response** | | | | |
| **Day 0** | | | | |
| n | 356 | 166 | 354 | 168 |
| GMT (95% CI) | 10.6 (10.1–11.1) | 10.0 (10.0–10.0) | 10.4 (10.0–10.9) | 10.1 (10.0–10.3) |
| **Day 35** | | | | |
| n | 355 | 168 | 351 | 168 |
| GMT (95% CI) | 1,302.7 (1,138.9–1,490.0) | 10.7 (10.0–11.5) | 898.9 (761.1–1061.6) | 10.8 (10.1–11.6) |
| GMFR (95% CI) | 123.8 (107.1–143.1) | 1.1 (1.0–1.2) | 86.1 (72.6–102.1) | 1.1 (1.0–1.1) |
| SCR*, % (95% CI) | 96.6 (94.1–98.2) | 2.4 (0.7–6.1) | 92.3 (89.0–94.9) | 2.4 (0.7–6.0) |
| **Serum IgG** | | | | |
| **Day 0** | | | | |
| n | 357 | 168 | 355 | 168 |
| GMEU, EU/mL (95% CI) | 117.0 (108.4–126.2) | 106.9 (101.8–112.1) | 121.7 (112.3–131.9) | 110.1 (104.3–116.2) |
| **Day 35** | | | | |
| n | 358 | 168 | 355 | 168 |
| GMEU, EU/mL (95% CI) | 64,414.3 (57,025.9–72,759.9) | 121.2 (107.1–137.2) | 37,775.6 (32,206.8–44,307.2) | 134.0 (116.1–154.8) |
| GMFR (95% CI) | 550.6 (477.5–634.8) | 1.1 (1.0–1.3) | 310.4 (260.2–370.3) | 1.2 (1.1–1.4) |
| SCR*, % (95% CI) | 97.8 (95.6–99.0) | 2.4 (0.7–6.0) | 95.5 (92.8–97.4) | 4.8 (2.1–9.2) |
| **hACE2 RBI** | | | | |
| **Day 0** | | | | |
| n | 357 | 168 | 355 | 169 |
| GMT (95% CI) | 5.2 (5.0–5.4) | 5.0 (5.0–5.0) | 5.1 (5.0–5.3) | 5.0 (5.0–5.0) |
| **Day 35** |  | | | |
| n | 358 | 168 | 354 | 169 |
| GMT (95% CI) | 222.4 (196.8–251.2) | 5.2 (5.0–5.4) | 135.7 (116.8–157.7) | 5.3 (5.0–5.7) |
| GMFR (95% CI) | 42.7 (37.5–48.6) | 1.0 (1.0–1.1) | 26.4 (22.7–30.7) | 1.1 (1.0–1.1) |
| SCR*, % (95% CI) | 91.6 (88.2–94.3) | 1.2 (0.1–4.2) | 82.5 (78.1–86.3) | 1.8 (0.4–5.1) |

^*^SCR was calculated based on the number of participants with a ≥4-fold increase in antibody concentration on Day 35 relative to Day 0 divided by the number of participants with a sample collected at Day 35. ELISA, enzyme-linked immunosorbent assay; EU, ELISA unit; GMEU, geometric mean ELISA unit; GMFR, geometric mean fold rise; GMT, geometric mean titer; hACE2, human angiotensin-converting enzyme 2; IgG, immunoglobulin G; nAb, neutralizing antibody; PP-IMM, per-protocol immunogenicity; RBI, receptor binding inhibition; SCR, seroconversion rate.

**Table S3.** Immunological responses to the ancestral strain after the 3^rd^ and 4^th^ doses of NVX‑CoV2373 in adults by age group (PP-IMM analysis sets).

|  | **Aged 18 to <65 years** | | | **Aged ≥65 years** | | |
| --- | --- | --- | --- | --- | --- | --- |
|  | **3^rd^ dose** | **4^th^ dose** | | **3^rd^ dose** | | **4^th^ dose** |
| **nAb response** | | | | | | |
| **n** | 199 | 91 | 24 | | 69 | |
| **Pre-dose** | | | | | | |
| GMT (95% CI) | 129.8 (104.7–161.0) | 1,960.9 (1,500.4–2,562.9) | | 84.8 (40.8–176.2) | | 1,280.0 (947.4–1,729.3) |
| **28 Days post dose** | | | | | | |
| GMT (95% CI) | 4,962.0 (4,298.4–5,728.0) | 5,278.4 (4,344.3–6,413.4) | | 4,974.2 (3,138.3–7,884.3) | | 4,023.1 (3,219.9–5,026.8) |
| GMFR (95% CI) | 38.2 (30.4–48.0) | 2.7 (2.2–3.3) | | 58.7 (27.9–123.5) | | 3.1 (2.5–3.9) |
| SCR*, % (95% CI) | 92.5 (87.9–95.7) | 38.5 (28.4–49.2) | | 91.7 (73.0–99.0) | | 47.8 (35.6–60.2) |
| **Serum IgG** | | | | | | |
| **n** | 200 | 91 | 25 | | 69 | |
| **Pre-dose** | | | | | | |
| GMEU, EU/mL  (95% CI) | 4,959.3  (4,114.7–5,977.2) | 67,064.3  (53,434.2–84,171.2) | | 3,502.7  (1,649.3–7,438.7) | | 50,214.6  (37,454.0–67,322.9) |
| **28 Days post dose** | | | | | | |
| GMEU, EU/mL  (95% CI) | 188,185.1  (164,988.7–214,642.8) | 163,480.1  (137,601.7–194,225.4) | | 119,676.1  (74,265.8–192,852.9) | | 122,256.4  (100,180.2–149,197.5) |
| GMFR (95% CI) | 37.9 (31.5–45.7) | 2.4 (2.1–2.8) | | 34.2 (16.9–69.1) | | 2.4 (2.0–2.9) |
| SCR*, % (95% CI) | 92.0 (87.3–95.4) | 19.8 (12.2–29.4) | | 84.0 (63.9–95.5) | | 26.1 (16.3–38.1) |
| **hACE2 RBI** | | | | | | |
| **n** | 201 | 91 | | 25 | | 69 |
| **Pre-dose** | | | | | | |
| GMT (95% CI) | 13.5 (11.2–16.4) | 181.0 (141.8–230.9) | | 11.4 (6.5–20.0) | | 131.3 (98.6–174.7) |
| **28 Days post dose** | | | | | | |
| GMT (95% CI) | 467.5 (405.5–538.9) | 427.7 (356.6–512.9) | | 372.3 (245.0–565.8) | | 336.3 (276.7–408.6) |
| GMFR (95% CI) | 34.6 (28.0–42.7) | 2.4 (2.0–2.8) | | 32.5 (15.7–67.5) | | 2.6 (2.1–3.1) |
| SCR*, % (95% CI) | 89.6 (84.5–93.4) | 22.0 (14.0–31.9) | | 84.0 (63.9–95.5) | | 24.6 (15.1–36.5) |

^*^For each dose, SCR was calculated based on the number of participants with a ≥4-fold increase in antibody concentration on Day 28 post dose relative to the respective Day 0 pre-dose, divided by the number of participants with a sample collected at Day 28 post dose.
ELISA, enzyme-linked immunosorbent assay; EU, ELISA unit; GMEU, geometric mean ELISA unit; GMFR, geometric mean fold rise; GMT, geometric mean titer; hACE2, human angiotensin-converting enzyme 2; IgG, immunoglobulin G; nAb, neutralizing antibody; PP-IMM, per-protocol immunogenicity; RBI, receptor binding inhibition; SCR, seroconversion rate.

**Table S4.** Solicited TEAEs^a^ (safety analysis sets).

|  | Adult main study | | | | | | Pediatric expansion (12 to <18 Years) | | | | | |
| --- | --- | --- | --- | --- | --- | --- | --- | --- | --- | --- | --- | --- |
| **Participants with TEAE, n (%)** | 1st dose | | 2nd dose | | 3rd dose | 4th dose | 1st dose | | 2nd dose | | 3rd dose | 4th dose |
|  | NVX-CoV2373  **(n=18,323)** | Placebo  **(n=9097)** | NVX-CoV2373  (n=17,345) | Placebo  (n=8439) | NVX-CoV2373  **(n=10,545)** | NVX-CoV2373  **(n=280)** | NVX-CoV2373  n = 1,448 | Placebo  (n=726) | NVX-CoV2373  (n=1,394) | Placebo  (n=686) | NVX-CoV2373 (n=1,251) | NVX-CoV2373 (n=163) |
| **Local** |  |  |  |  |  |  |  |  |  |  |  |  |
| **Any local** | 10,609 (57.9) | 1,927 (21.2) | 13,670 (78.8) | 1823 (21.6) | 8332 (79.0) | 200 (71.4) | 948 (65.5) | 207 (28.5) | 1,050 (75.3) | 141 (20.6) | 969 (77.5) | 122 (74.8) |
| Grade 3 | 194 (1.1) | 21 (0.2) | 1134 (6.5) | 24 (0.3) | 1014 (9.6) | 11 (3.9) | 22 (1.5) | 4 (0.6) | 118 (8.5) | 4 (0.6) | 167 (13.3) | 18 (11.0) |
| Grade 4 | 0 | 0 | 2 (<0.1) | 0 | 9 (<0.1) | 0 | 0 | 0 | 0 | 0 | 1 (<0.1) | 1 (0.6) |
| **Pain** | 6279 (34.3) | 1014 (11.1) | 10,327 (59.5) | 1154 (13.7) | 6453 (61.2) | 137 (48.9) | 647 (44.7) | 126 (17.4) | 850 (61.0) | 102 (14.9) | 812 (64.9) | 106 (65.0) |
| Grade 3 | 55 (0.3) | 3 (<0.1) | 301 (1.7) | 7 (<0.1) | 312 (3.0) | 1 (0.4) | 10 (0.7) | 2 (0.3) | 38 (2.7) | 3 (0.4) | 61 (4.9) | 5 (3.1) |
| Grade 4 | 0 | 0 | 1 (<0.1) | 0 | 6 (<0.1) | 0 | 0 | 0 | 0 | 0 | 0 | 1 (0.6) |
| **Tenderness** | 9569 (52.2) | 1528 (16.8) | 12,716 (73.3) | 1329 (15.7) | 7698 (73.0) | 187 (66.8) | 819 (56.6) | 153 (21.1) | 909 (65.2) | 97 (14.1) | 828 (66.2) | 102 (62.6) |
| Grade 3 | 156 (0.9) | 18 (0.2) | 844 (4.9) | 18 (0.2) | 711 (6.7) | 8 (2.9) | 16 (1.1) | 2 (0.3) | 93 (6.7) | 1 (0.1) | 116 (9.3) | 12 (7.4) |
| Grade 4 | 0 | 0 | 2 (<0.1) | 0 | 7 (<0.1) | 0 | 0 | 0 | 0 | 0 | 1 (<0.1) | 1 (0.6) |
| **Redness** | 165 (0.9) | 28 (0.3) | 1142 (6.6) | 30 (0.4) | 953 (9.0) | 31 (11.1) | 15 (1.0) | 5 (0.7) | 104 (7.5) | 0 | 129 (10.3) | 13 (8.0) |
| Grade 3 | 2 (<0.1) | 0 | 126 (0.7) | 2 (<0.1) | 183 (1.7) | 2 (0.7) | 0 | 0 | 10 (0.7) | 0 | 29 (2.3) | 3 (1.8) |
| Grade 4 | 0 | 0 | 0 | 0 | 0 | 0 | 0 | 0 | 0 | 0 | 0 | 0 |
| **Swelling** | 153 (0.8) | 25 (0.3) | 1062 (6.1) | 26 (0.3) | 873 (8.3) | 27 (9.6) | 20 (1.4) | 3 (0.4) | 111 (8.0) | 1 (0.1) | 119 (9.5) | 9 (5.5) |
| Grade 3 | 4 (<0.1) | 1 (<0.1) | 72 (0.4) | 1 (<0.1) | 109 (1.0) | 0 | 0 | 0 | 8 (0.6) | 0 | 18 (1.4) | 1 (0.6) |
| Grade 4 | 0 | 0 | 0 | 0 | 0 | 0 | 0 | 0 | 0 | 0 | 0 | 0 |
| **Systemic** |  |  |  |  |  |  |  |  |  |  |  |  |
| **Any systemic** | 8725 (47.6) | 3638 (40.0) | 12,044 (69.4) | 3026 (35.9) | 7607 (72.1) | 171 (61.1) | 799 (55.2) | 296 (40.8) | 1,038 (74.5) | 198 (28.9) | 1,013 (81.0) | 132 (81.0) |
| Grade 3 | 425 (2.3) | 187 (2.1) | 2080 (12.0) | 171 (2.0) | 1794 (17.0) | 18 (6.4) | 52 (3.6) | 25 (3.4) | 304 (21.8) | 23 (3.4) | 371 (29.7) | 43 (26.4) |
| Grade 4 | 12 (<0.1) | 3 (<0.1) | 10 (<0.1) | 5 (<0.1) | 23 (0.2) | 0 | 0 | 0 | 2 (0.1) | 0 | 5 (0.4) | 1 (0.6) |
| **Fatigue** | 4685 (25.6) | 2031 (22.3) | 8565 (49.4) | 1848 (21.9) | 5543 (52.6) | 127 (45.4) | 350 (24.2) | 112 (15.4) | 695 (49.9) | 100 (14.6) | 717 (57.3) | 87 (53.4) |
| Grade 3 | 227 (1.2) | 103 (1.1) | 1432 (8.3) | 110 (1.3) | 1287 (12.2) | 13 (4.6) | 23 (1.6) | 9 (1.2) | 185 (13.3) | 10 (1.5) | 210 (16.8) | 24 (14.7) |
| Grade 4 | 3 (<0.1) | 1 (<0.1) | 2 (<0.1) | 3 (<0.1) | 12 (0.1) | 0 | 0 | 0 | 0 | 0 | 1 (<0.1) | 0 |
| **Headache** | 4560 (24.9) | 2072 (22.8) | 7704 (44.4) | 1654 (19.6) | 4803 (45.5) | 89 (31.8) | 440 (30.4) | 181 (24.9) | 793 (56.9) | 119 (17.3) | 790 (63.1) | 95 (58.3) |
| Grade 3 | 146 (0.8) | 62 (0.7) | 518 (3.0) | 38 (0.5) | 569 (5.4) | 7 (2.5) | 13 (0.9) | 12 (1.7) | 87 (6.2) | 14 (2.0) | 154 (12.3) | 16 (9.8) |
| Grade 4 | 4 (<0.1) | 1 (<0.1) | 3 (<0.1) | 2 (<0.1) | 6 (< 0.1) | 0 | 0 | 0 | 1 (<0.1) | 0 | 2 (0.2) | 0 |
| **Muscle pain** | 4153 (22.7) | 1211 (13.3) | 8332 (48.0) | 1015 (12.0) | 5440 (51.6) | 115 (41.1) | 492 (34.0) | 114 (15.7) | 683 (49.0) | 82 (12.0) | 754 (60.3) | 100 (61.3) |
| Grade 3 | 82 (0.4) | 35 (0.4) | 850 (4.9) | 30 (0.4) | 893 (8.5) | 7 (2.5) | 17 (1.2) | 4 (0.6) | 104 (7.5) | 6 (0.9) | 143 (11.4) | 14 (8.6) |
| Grade 4 | 2 (<0.1) | 1 (<0.1) | 2 (<0.1) | 4 (<0.1) | 11 (0.1) | 0 | 0 | 0 | 0 | 0 | 1 (<0.1) | 0 |
| **Malaise** | 2692 (14.7) | 1063 (11.7) | 6742 (38.9) | 1038 (12.3) | 4312 (40.9) | 100 (35.7) | 215 (14.8) | 67 (9.2) | 560 (40.2) | 51 (7.4) | 566 (45.2) | 83 (50.9) |
| Grade 3 | 139 (0.8) | 56 (0.6) | 1083 (6.2) | 59 (0.7) | 985 (9.3) | 5 (1.8) | 16 (1.1) | 7 (1.0) | 126 (9.0) | 4 (0.6) | 170 (13.6) | 17 (10.4) |
| Grade 4 | 6 (<0.1) | 1 (<0.1) | 5 (<0.1) | 2 (<0.1) | 12 (0.1) | 0 | 0 | 0 | 0 | 0 | 1 (<0.1) | 0 |
| **Joint pain** | 1414 (7.7) | 599 (6.6) | 3855 (22.2) | 577 (6.8) | 2768 (26.2) | 57 (20.4) | 102 (7.0) | 35 (4.8) | 225 (16.1) | 21 (3.1) | 275 (22.0) | 29 (17.8) |
| Grade 3 | 53 (0.3) | 29 (0.3) | 419 (2.4) | 24 (0.3) | 522 (5.0) | 6 (2.1) | 6 (0.4) | 1 (0.1) | 40 (2.9) | 2 (0.3) | 50 (4.0) | 7 (4.3) |
| Grade 4 | 1 (<0.1) | 0 | 2 (<0.1) | 2 (<0.1) | 6 (< 0.1) | 0 | 0 | 0 | 0 | 0 | 1 (<0.1) | 0 |
| **Nausea/vomiting** | 1168 (6.4) | 504 (5.5) | 1949 (11.2) | 456 (5.4) | 1255 (11.9) | 18 (6.4) | 113 (7.8) | 55 (7.6) | 277 (19.9) | 33 (4.8) | 292 (23.3) | 37 (22.7) |
| Grade 3 | 18 (<0.1) | 7 (<0.1) | 31 (0.2) | 7 (<0.1) | 47 (0.4) | 0 | 2 (0.1) | 3 (0.4) | 14 (1.0) | 3 (0.4) | 20 (1.6) | 3 (1.8) |
| Grade 4 | 4 (<0.1) | 2 (<0.1) | 5 (<0.1) | 2 (<0.1) | 5 (< 0.1) | 0 | 0 | 0 | 1 (<0.1) | 0 | 0 | 0 |
| **Fever** | 64 (0.3) | 33 (0.4) | 977 (5.6) | 23 (0.3) | 811 (7.7) | 13 (4.6) | 9 (0.6) | 4 (0.6) | 236 (16.9) | 1 (0.1) | 211 (16.9) | 18 (11.0) |
| Grade 3 | 8 (<0.1) | 7 (<0.1) | 63 (0.4) | 3 (<0.1) | 105 (1.0) | 0 | 1 (<0.1) | 0 | 32 (2.3) | 0 | 44 (3.5) | 4 (2.5) |
| Grade 4 | 3 (<0.1) | 0 | 0 | 0 | 5 (< 0.1) | 0 | 0 | 0 | 0 | 0 | 3 (0.2) | 1 (0.6) |

^a^Reactogenicity data were collected from participants who had at least one eDiary entry, reported here based on the following data cut dates: adults, December 20, 2023; adolescents: August 6, 2022 (primary series and 3^rd^ dose); December 27, 2023 (4^th^ dose). TEAE, treatment-emergent adverse event.

**Table S5.** Duration of solicited TEAEs within the reactogenicity period^a^ (safety analysis sets).

|  | Adult main study | | | | | | Pediatric expansion (12 to <18 Years) | | | | | | | | |
| --- | --- | --- | --- | --- | --- | --- | --- | --- | --- | --- | --- | --- | --- | --- | --- |
|  | 1st dose | | 2nd dose | | 3rd dose | 4th dose | 1st dose | | | 2nd dose | | | 3rd dose | | 4th dose |
|  | NVX-CoV2373  **(n=18,323)** | Placebo  **(n=9097)** | NVX-CoV2373  (n=17,345) | Placebo  (n=8439) | NVX-CoV2373  **(n=10,545)** | NVX-CoV2373  **(n=280)** | NVX-CoV2373  (n=1,448) | Placebo  (n=726) | | NVX-CoV2373  (n=1,394) | | Placebo  (n=686) | NVX-CoV2373 (n=1,251) | | NVX-CoV2373 (n=163) |
| **Local TEAEs** |  | | | | | |  | | | | | | | | |
| **Any local** |  | | | | | |  | | | | | | | | |
| Median, days (range) | 2 (1–7) | 1 (1–7) | 3 (1–7) | 1 (1–7) | 3 (1–7) | 3 (1–7) | 2 (1–7) | 1 (1–7) | | 3 (1–7) | | 1 (1–7) | 3 (1–7) | | 2 (1–7) |
| Persisted >7 days, n | 8 | 2 | 29 | 1 | 27 | 0 | 5 | 0 | | 2 | | 1 | 2 | | 0 |
| **Pain** |  | | | | | |  | | | | | | | | |
| Median, days (range) | 1 (1–7) | 1 (1–7) | 2 (1–7) | 1 (1–7) | 2 (1–7) | 2 (1–7) | 2 (1–7) | 1 (1–7) | | 2 (1–7) | | 1 (1–7) | 2 (1–7) | | 2 (1–7) |
| Persisted >7 days, n | 1 | 1 | 6 | 0 | 6 | 0 | 3 | 0 | | 2 | | 1 | 0 | | 0 |
| **Tenderness** |  | | | | | |  | | | | | | | | |
| Median, days (range) | 2 (1–7) | 1 (1–7) | 3 (1–7) | 1 (1–7) | 3 (1–7) | 3 (1–7) | 2 (1–7) | 1 (1–7) | | 2 (1–7) | | 1 (1–7) | 2 (1–7) | | 2.5 (1–7) |
| Persisted >7 days, n | 5 | 2 | 16 | 1 | 7 | 0 | 3 | 0 | | 2 | | 1 | 0 | | 0 |
| **Redness** |  | | | | | |  | | | | | | | | |
| Median, days (range) | 1 (1–7) | 1 (1–5) | 2 (1–7) | 1 (1–6) | 2 (1–7) | 2 (1–5) | 2 (1–6) | 1 (1–1) | | 2 (1–6) | | 0 | 2 (1–6) | | 2 (1–4) |
| Persisted >7 days, n | 1 | 0 | 9 | 1 | 12 | 0 | 0 | 0 | | 0 | | 0 | 2 | | 0 |
| **Swelling** |  | | | | | |  | | | | | | | | |
| Median, days (range) | 1 (1–7) | 1 (1–6) | 2 (1–7) | 1 (1–5) | 2 (1–7) | 2 (1–5) | 1 (1–6) | 2 (1–3) | | 2 (1–6) | | 1 (1–1) | 2 (1–6) | | 3 (1–5) |
| Persisted >7 days, n | 1 | 0 | 6 | 0 | 11 | 0 | 0 | 0 | | 0 | | 0 | 1 | | 0 |
| **Systemic TEAEs** |  | | | | | |  | | | | | | | | |
| **Any systemic** |  | | | | | |  | | | | | | | | |
| Median, days (range) | 2 (1–7) | 2 (1–7) | 2 (1–7) | 2 (1–7) | 2 (1–7) | 2 (1–7) | 2 (1–7) | 1 (1–7) | | 2 (1–7) | | 1 (1–7) | 2 (1–7) | | 2 (1–7) |
| Persisted >7 days, n | 41 | 12 | 35 | 20 | 16 | 0 | 12 | 4 | | 8 | | 3 | 1 | | 0 |
| **Fatigue** |  | | | | | |  | | | | | | | | |
| Median, days (range) | 1 (1–7) | 1 (1–7) | 1 (1–7) | 1 (1–7) | 1 (1–7) | 2 (1–7) | 1 (1–7) | | 1 (1–7) | 1 (1–7) | 1 (1–7) | | 1 (1–7) | 1 (1–6) | |
| Persisted >7 days, n | 10 | 2 | 8 | 5 | 4 | 0 | 6 | | 0 | 2 | 2 | | 1 | 0 | |
| **Headache** |  | | | | | |  | | | | | | | | |
| Median, days (range) | 1 (1–7) | 1 (1–7) | 1 (1–7) | 1 (1–7) | 1 (1–7) | 1 (1–7) | 1 (1–7) | | 1 (1–7) | 1 (1–7) | 1 (1–7) | | 2 (1–7) | 2 (1–7) | |
| Persisted >7 days, n | 16 | 7 | 12 | 13 | 10 | 0 | 7 | | 1 | 5 | 1 | | 1 | 0 | |
| **Muscle pain** |  | | | | | |  | | | | | | | | |
| Median, days (range) | 1 (1–7) | 1 (1–7) | 1 (1–7) | 1 (1–7) | 2 (1–7) | 2 (1–7) | 1 (1–7) | | 1 (1–7) | 2 (1–7) | 1 (1–7) | | 2 (1–7) | 2 (1–7) | |
| Persisted >7 days, n | 7 | 2 | 7 | 4 | 0 | 0 | 2 | | 2 | 3 | 0 | | 1 | 0 | |
| **Malaise** |  | | | | | |  | | | | | | | | |
| Median, days (range) | 1 (1–7) | 1 (1–7) | 1 (1–7) | 1 (1–7) | 1 (1–7) | 1 (1–7) | 1 (1–7) | | 1 (1–7) | 1 (1–7) | 1 (1–7) | | 1 (1–7) | 1 (1–7) | |
| Persisted >7 days, n | 1 | 0 | 2 | 0 | 1 | 0 | 1 | | 1 | 1 | 1 | | 0 | 0 | |
| **Joint pain** |  | | | | | |  | | | | | | | | |
| Median, days (range) | 1 (1–7) | 1 (1–7) | 1 (1–7) | 1 (1–7) | 1 (1–7) | 1 (1–6) | 1 (1–7) | | 1 (1–5) | 1 (1–6) | 1 (1–7) | | 1 (1–7) | 1 (1–6) | |
| Persisted >7 days, n | 5 | 2 | 7 | 5 | 2 | 0 | 2 | | 1 | 3 | 0 | | 0 | 0 | |
| **Nausea/vomiting** |  | | | | | |  | | | | | | | | |
| Median, days (range) | 1 (1–7) | 1 (1–7) | 1 (1–7) | 1 (1–7) | 1 (1–7) | 1 (1–7) | 1 (1–7) | | 1 (1–7) | 1 (1–7) | 1 (1–7) | | 1 (1–7) | 1 (1–6) | |
| Persisted >7 days, n | 8 | 0 | 7 | 1 | 1 | 0 | 0 | | 0 | 0 | 1 | | 1 | 0 | |
| **Fever** |  | | | | | |  | | | | | | | | |
| Median, days (range) | 1 (1–7) | 1 (1–3) | 1 (1–6) | 1 (1–5) | 1 (1–6) | 1 (1–6) | 1 (1–3) | | 1 (1–1) | 1 (1–2) | 1 (1–1) | | 1 (1–4) | 1 (1–2) | |
| Persisted >7 days, n | 1 | 1 | 1 | 0 | 2 | 0 | 0 | | 0 | 2 | 0 | | 1 | 0 | |

^a^Reactogenicity data were collected from participants who had at least one eDiary entry, reported here based on the following data cut dates: adults, December 20, 2023; adolescents: August 6, 2022 (primary series and 3^rd^ dose); December 27, 2023 (4^th^ dose). TEAE, treatment-emergent adverse event.

**Table S6.** Overall summary of unsolicited TEAEs (safety analysis sets).

|  | Adult main study | | | | Pediatric expansion (12 to <18 years) | | | |
| --- | --- | --- | --- | --- | --- | --- | --- | --- |
| **Participants, n (%)** | Primary series | | 3^rd^ dose | 4^th^ dose | Primary series | | 3^rd^ dose | 4^th^ dose |
|  | NVX-CoV2373  **(n=19,735)** | Placebo  **(n=9847)** | NVX-CoV2373  **(n=13,354)** | NVX-CoV2373  **(n=359)** | NVX-CoV2373 (n=1,487) | Placebo (n=745) | NVX-CoV2373 (n=1,499) | NVX-CoV2373 (n=205) |
|  | *Through 28 days after series/dose* | | | | *Through 28 days after series/dose* | | | |
| Any | 2389 (12.1) | 1139 (11.6) | 669 (5.0) | 7 (1.9) | 235 (15.8) | 119 (16.0) | 87 (5.8) | 7 (3.4) |
| Related | 480 (2.4) | 146 (1.5) | 100 (0.7) | 0 | 42 (2.8) | 8 (1.1) | 17 (1.1) | 1 (0.5) |
| Severe | 153 (0.8) | 65 (0.7) | 48 (0.4) | 0 | 6 (0.4) | 2 (0.3) | 9 (0.6) | 0 |
| Related | 20 (0.1) | 6 (<0.1) | 11 (<0.1) | 0 | 0 | 0 | 1 (<0.1) | 0 |
| MAAE | 1006 (5.1) | 498 (5.1) | 379 (2.8) | 5 (1.4) | – | – | – | 6 (2.9) |
|  | *TEAE through end of study^a^* | | | | *TEAE through data cutoff date^a^* | | | |
| MAAE | 1138 (5.8) | 560 (5.7) | 681 (5.1) | 6 (1.7) | 95 (6.4) | 51 (6.8) | 43 (2.9) | NA |
| Related | 103 (0.5) | 28 (0.3) | 35 (0.3) | 0 | 5 (0.3) | 3 (0.4) | 3 (0.2) | 0 |
| SAE | 238 (1.2) | 129 (1.3) | 397 (3.0) | 4 (1.1) | 7 (0.5) | 2 (0.3) | 9 (0.6) | 3 (1.5) |
| Related | 6 (<0.1) | 3 (<0.1) | 5 (<0.1) | 0 | 0 | 0 | 0 | 0 |
| PIMMC | 27 (0.1) | 13 (0.1) | 24 (0.2) | 0 | 1 (<0.1) | 0 | 0 | 0 |
| Related | 17 (<0.1) | 3 (<0.1) | 2 (<0.1) | 0 | 0 | 0 | 0 | 0 |
| AESI relevant to COVID-19 | 6 (<0.1) | 5 (<0.1) | 9 (<0.1) | 0 | 0 | 0 | 0 | 0 |
| Related | 0 | 1 (<0.1) | 0 | 0 | 0 | 0 | 0 | 0 |
| Leading to study vaccine discontinuation | 54 (0.3) | 18 (0.2) | – | – | 1 (<0.1) | 1 (0.1) | 0 | NA |
| Related | 12 (<0.1) | 3 (<0.1) | – | – | 0 | 0 | 0 | NA |
| Leading to study discontinuation | 28 (0.1) | 13 (0.1) | 27 (0.2) | 1 (0.3) | 0 | 0 | 0 | NA |
| Related | 4 (<0.1) | 2 (<0.1) | 0 | 0 | 0 | 0 | 0 | NA |
| Deaths | 12 (<0.1) | 9 (<0.1) | 25 (0.2) | 1 (0.3) | 0 | 0 | 0 | 0 |

^a^Adults: December 20, 2023. Adolescents: August 6, 2022 (primary series and 3^rd^ dose); December 27, 2023 (4^th^ dose). Not all details for the pediatric population after the 4^th^ dose are available as these analyses are still ongoing.
AE, adverse event; AESI, adverse event of special interest; COVID-19, coronavirus disease 2019; MAAE, medically attended adverse event; NA, not available; PIMMC, potential immune-mediated medical condition; SAE, serious adverse event; TEAE, treatment-emergent adverse event.

**Figure S1.** Study design.

Timing of study doses and assessments are shown. After accrual of safety data required for emergency use authorization, participants could opt to continue in a blinded crossover and receive alternate treatment from their original randomization group. Actual median time between doses (2^nd^ and 3^rd^, 3^rd^ and 4^th^) are shown for adults and adolescents.
*Blood samples for immunogenicity were collected before administration of each study dose.
^†^Solicited TEAEs (reactogenicity) were recorded in an eDiary for 7 days after each study dose.
^‡^Unsolicited TEAEs, MAAEs, AESI, and SAEs were documented through 28 days after each study dose. AESI, SAEs, and related MAAEs were collected through end of study.
^§^Timing comprises participants in initial primary series group, not including the crossover group.
AESI, adverse events of special interest; EOS, end of study; MAAE, medically attended adverse event; SAE, serious adverse event; TEAE, treatment-emergent adverse event.

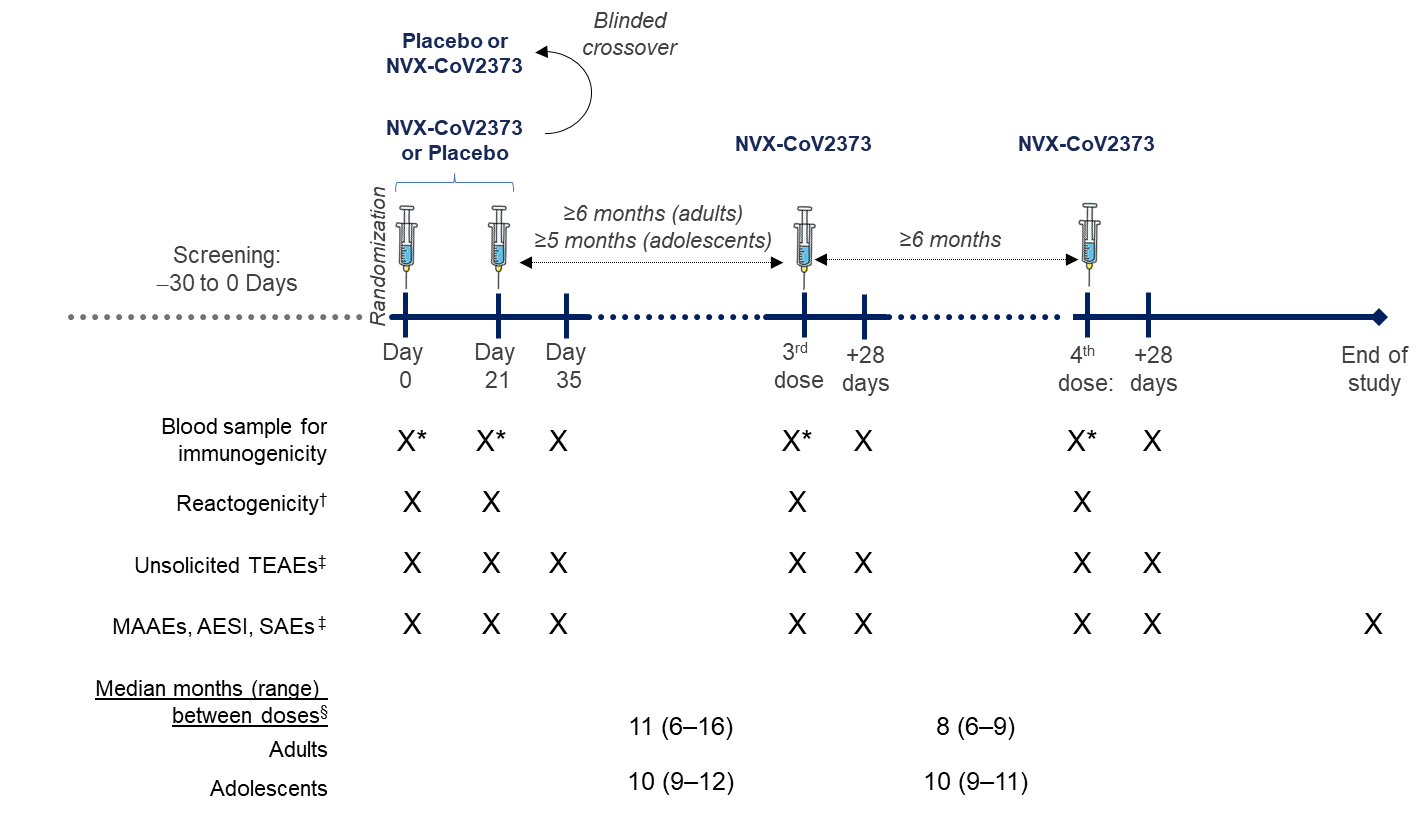

**Figure S2.** Anti-rS IgG responses against the ancestral strain after each study dose (PP-IMM analysis sets).

(A) Adult main study. (B) Pediatric expansion adolescent population.

Baseline is defined as the last available assessment before administration of trial vaccine for the 1st, 3rd, or 4th dose. Post-dose is 14 days (Day 35) after the primary series and 28 days after the 3^rd^ or 4^th^ dose. GMFRs were calculated based on titer comparison of respective post-dose and baseline samples for each group. SCR was calculated based on the number of participants with a ≥4-fold increase in antibody concentration from baseline divided by the number of participants with a sample collected at the designated post-vaccination time. GMEU, geometric mean ELISA unit; GMFR, geometric mean fold rise; IgG, immunoglobulin G; PP-IMM, per-protocol immunogenicity; rS, recombinant spike; SCR, seroconversion rate.

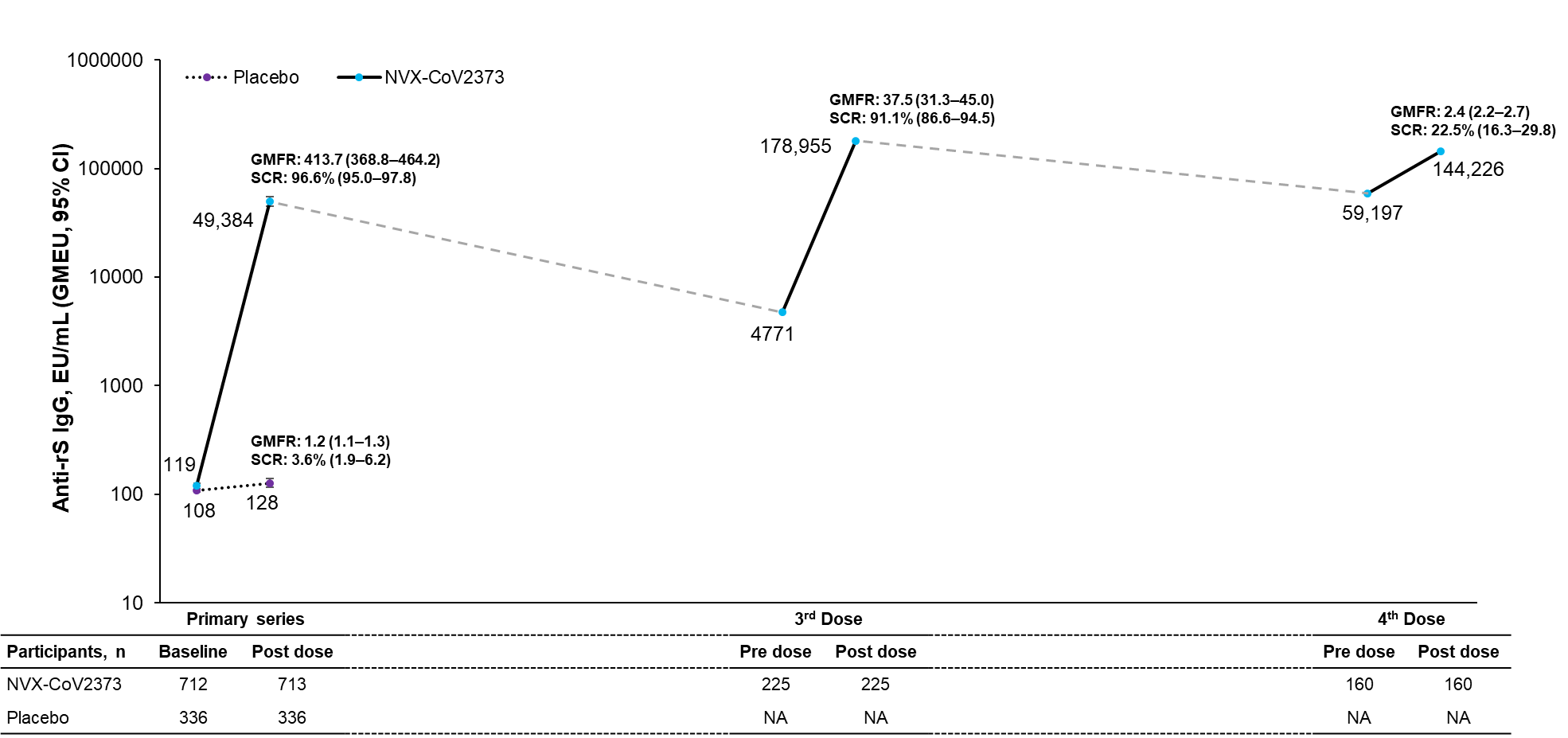
**A**

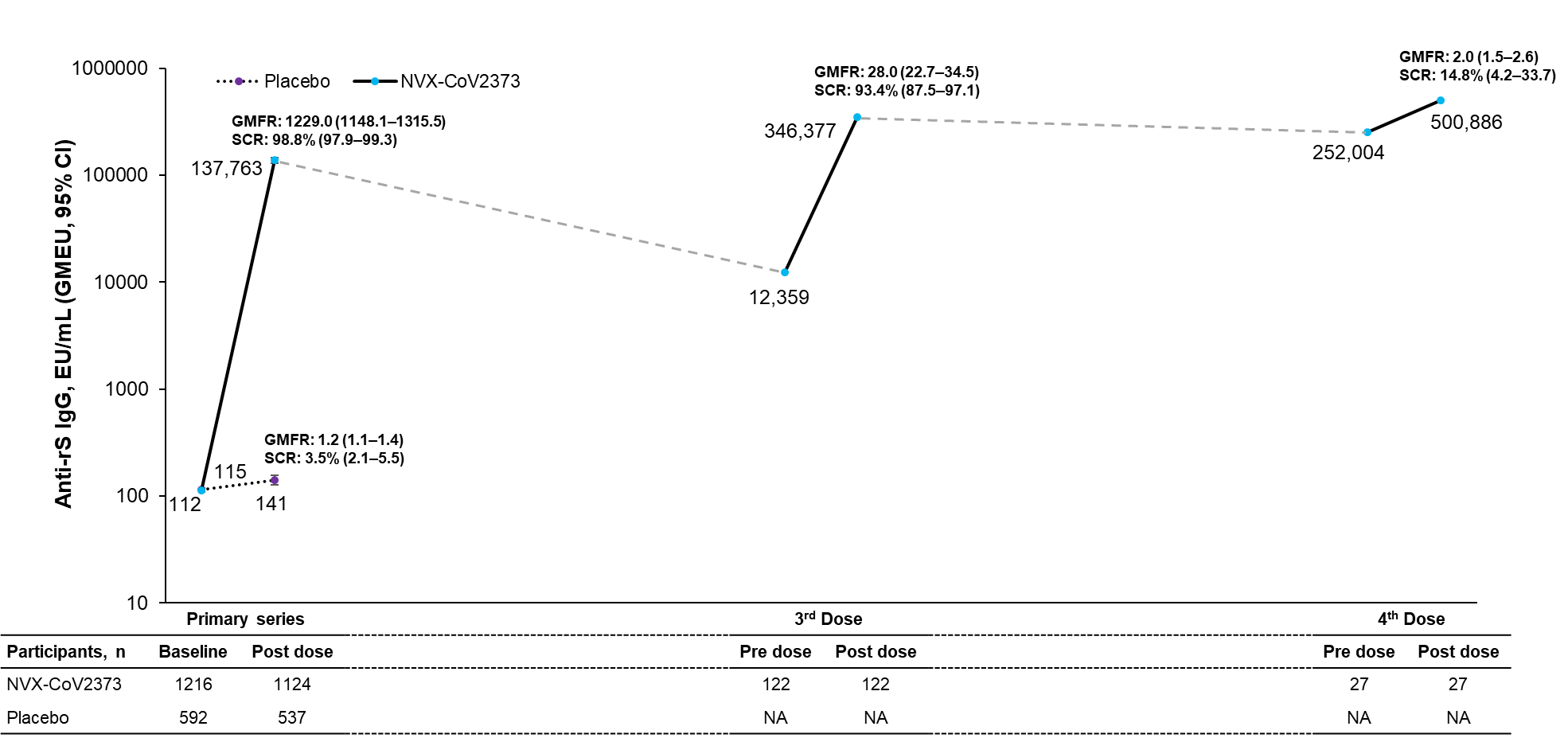
**B**

**Figure S3.** hACE2 RBI responses against the ancestral strain and after each study dose (PP-IMM analysis sets).

(A) Adult main study. (B) Pediatric expansion adolescent population.

Baseline is defined as the last available assessment before administration of trial vaccine for the 1st, 3rd, or 4th dose. Post-dose is 14 days (Day 35) after the primary series and 28 days after the 3^rd^ or 4^th^ dose. GMFRs were calculated based on titer comparison of respective post-dose and baseline samples for each group. SCR was calculated based on the number of participants with a ≥4-fold increase in antibody concentration from baseline divided by the number of participants with a sample collected at the designated post-vaccination time. GMFR, geometric mean fold rise; GMT, geometric mean titer; hACE2, human angiotensin-converting enzyme 2; IgG, immunoglobulin G; PP-IMM, per-protocol immunogenicity; RBI, receptor binding inhibition; SCR, seroconversion rate.

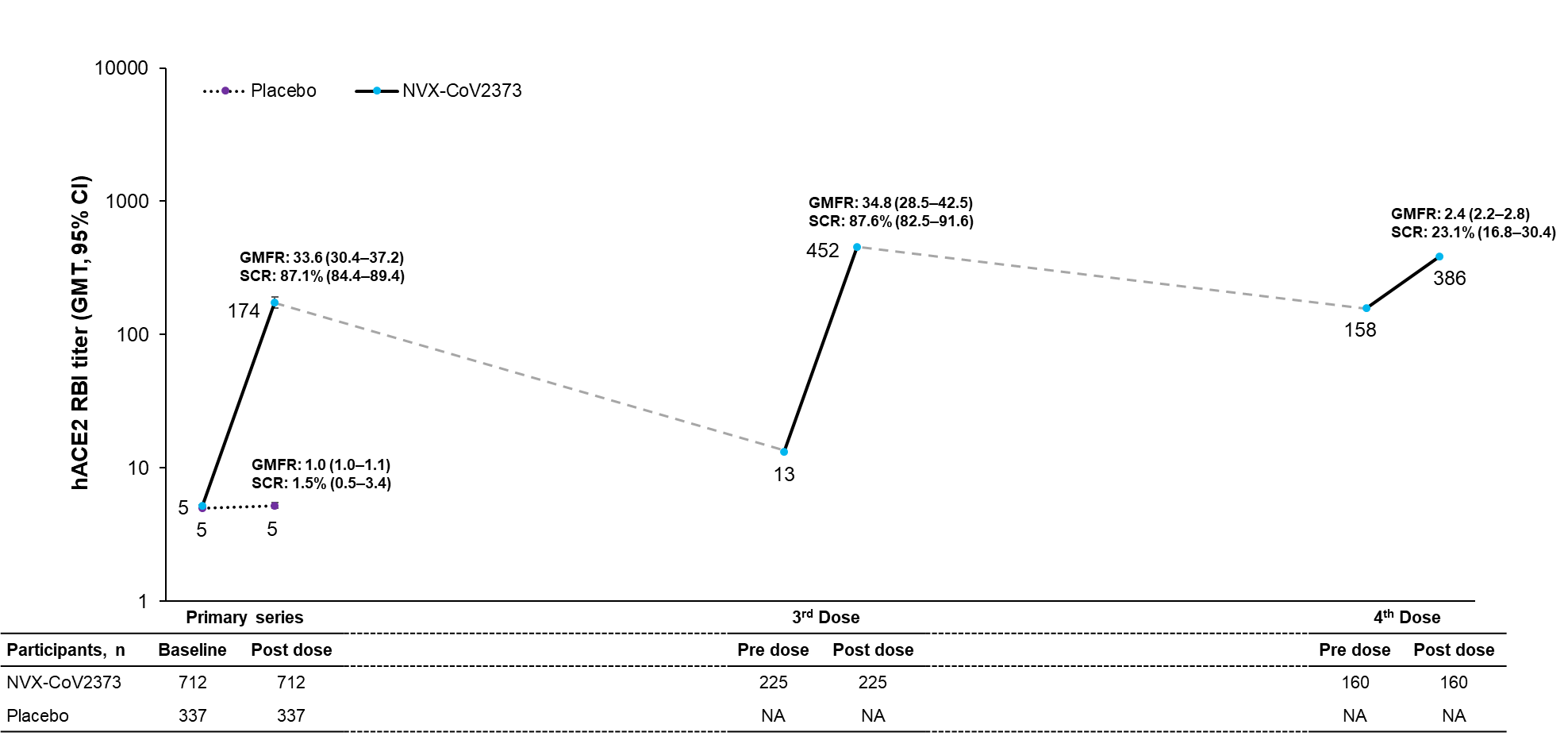
**A**

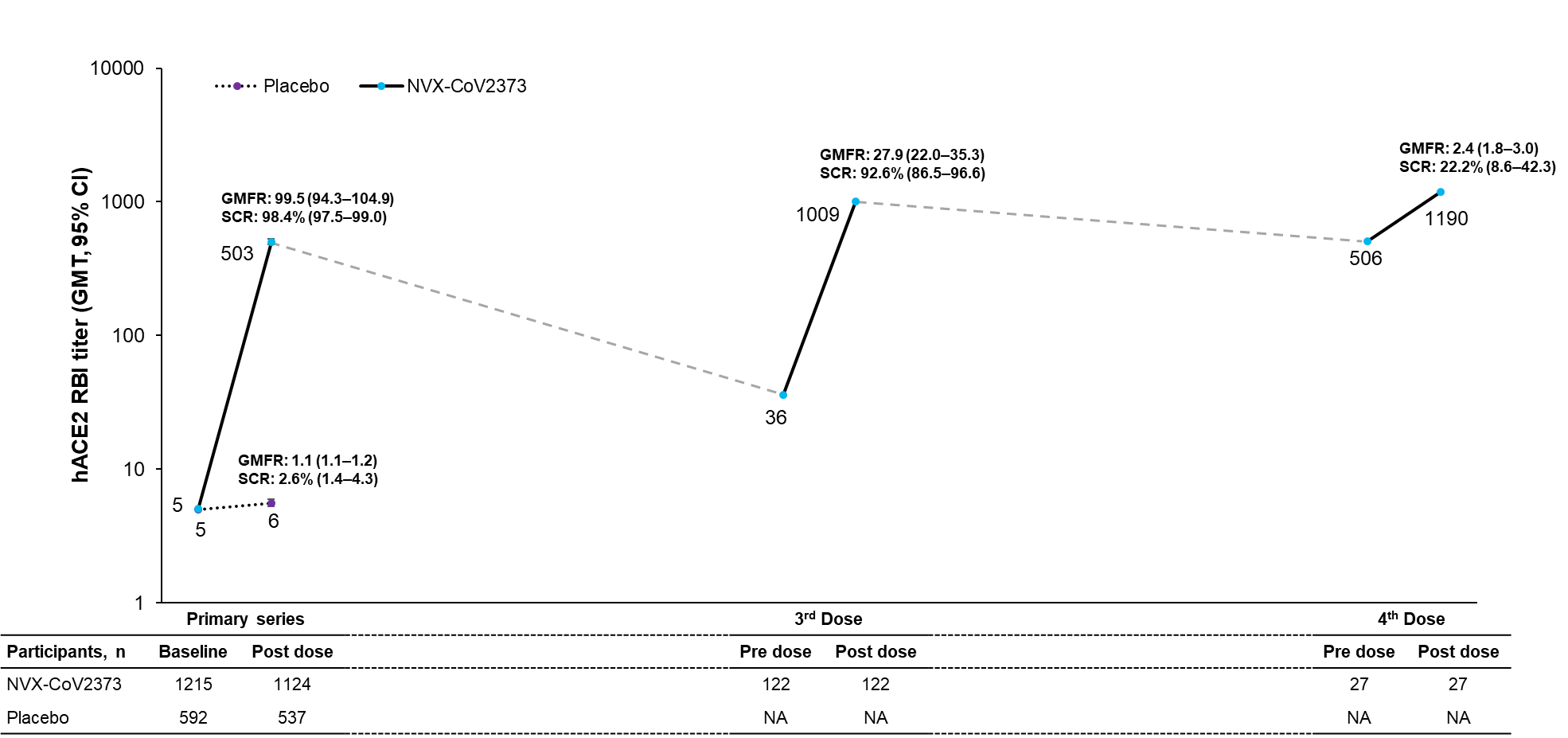

**B**

**Figure S4.** Primary series responses against the ancestral strain in the adult main study, regardless of baseline serostatus (PP-IMM-2 analysis sets).

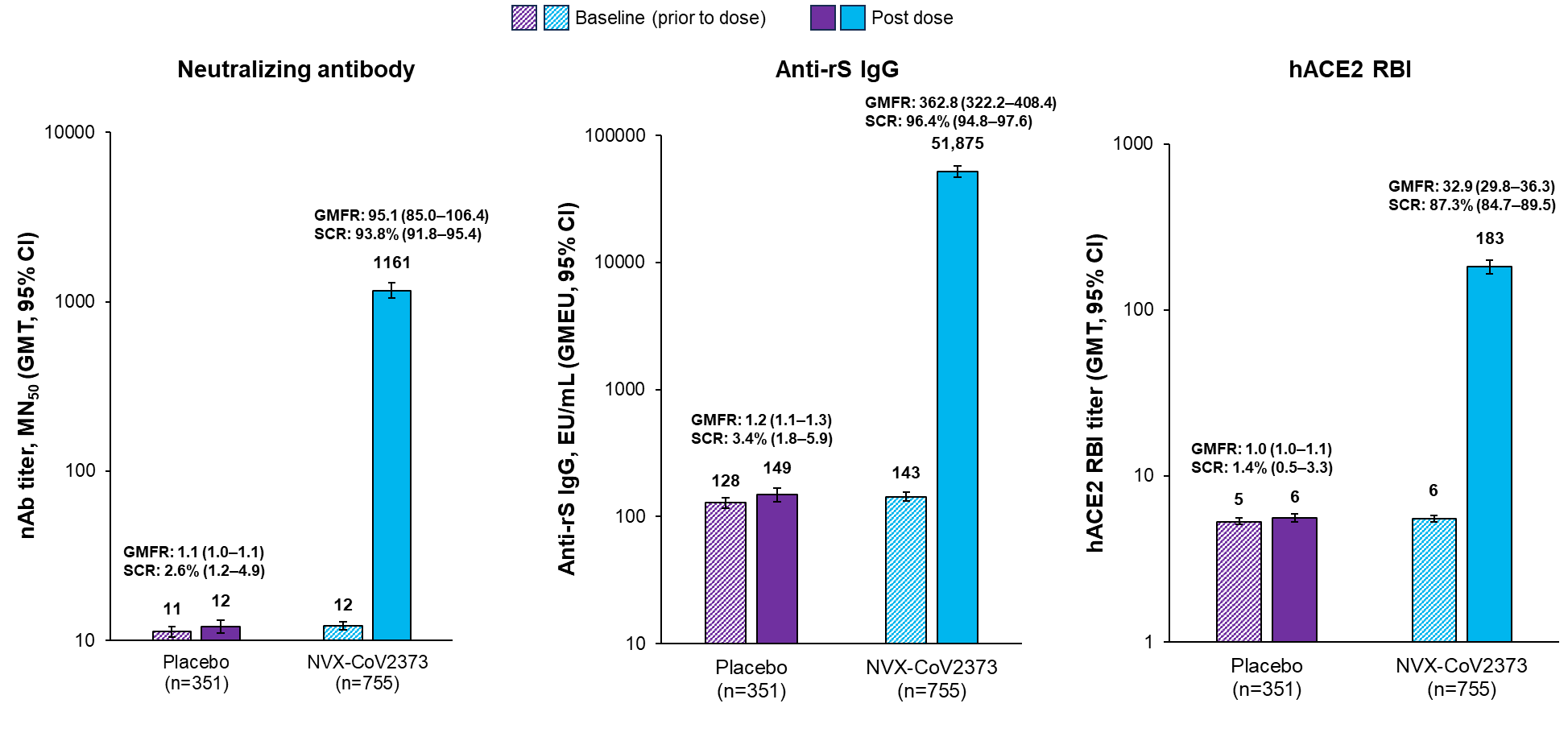

**Figure S5.** Durability of immunogenic responses 4 months (120 days) and 8 months (240 days) after the 3^rd^ dose of NVX-CoV2373 (PP-IMM subset).

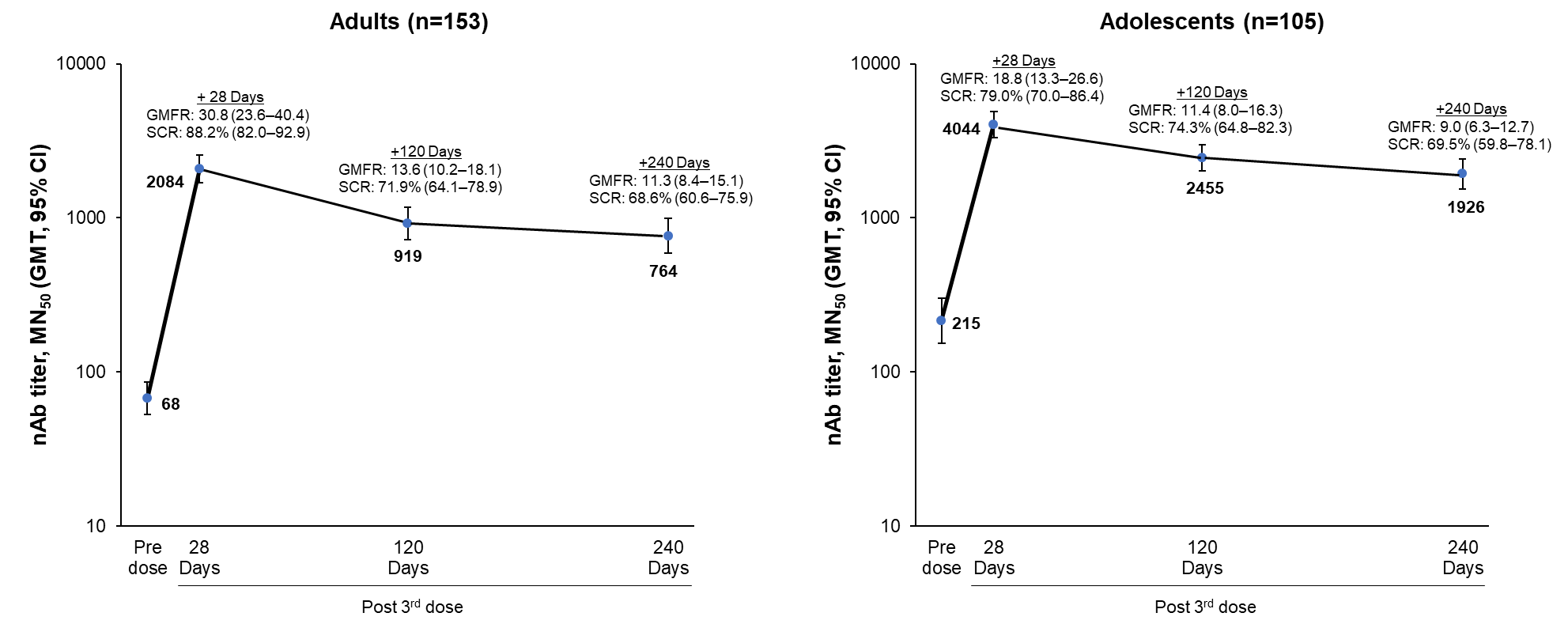
**A. nAb responses**

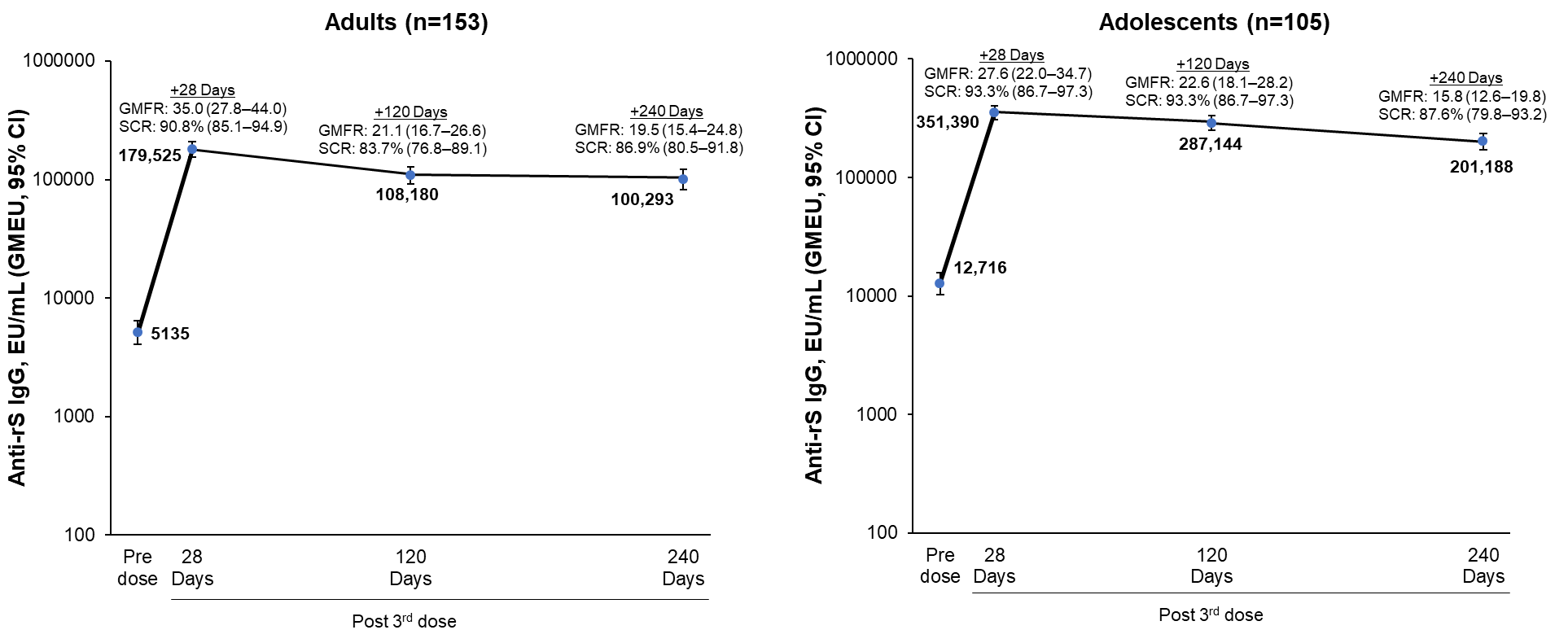
**B. Anti-rS IgG responses**

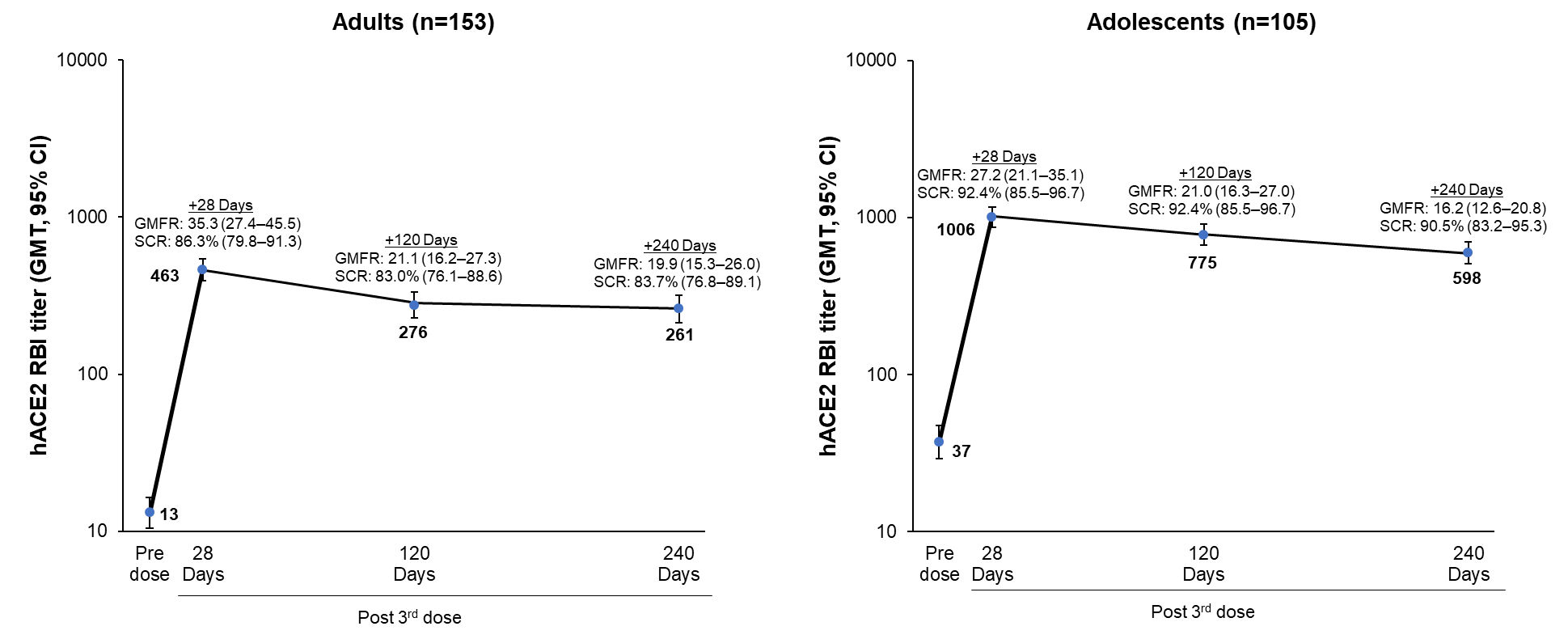
**C. hACE2 RBI**
